# The impact of heating, ventilation and air conditioning (HVAC) design features on the transmission of viruses, including SARS-CoV-2: an overview of reviews

**DOI:** 10.1101/2021.09.22.21263515

**Authors:** Gail M. Thornton, Emily Kroeker, Brian A. Fleck, Lexuan Zhong, Lisa Hartling

## Abstract

**Background:** The 2019 novel coronavirus or severe acute respiratory syndrome coronavirus 2 (SARS-CoV-2) outbreak was declared a pandemic by the World Health Organization (WHO) in March 2020. Almost two years later (early-February 2022), the WHO reported over 383 million cases of the disease caused by the virus with over 5.6 million deaths worldwide. Debate regarding routes of transmission was substantial early in the pandemic; however, airborne transmission emerged as an important consideration. Infectious airborne agents can spread within the built environment through heating, ventilation, and air conditioning (HVAC) systems. Multiple features of HVAC systems can influence transmission (e.g., ventilation, filtration, ultraviolet radiation, humidity). Understanding how HVAC features influence airborne transmission is critical to mitigate the spread of infectious agents.

**Objective:** Given airborne transmission of SARS-CoV-2, an overview of reviews was conducted to understand what is already known from the scientific literature about how virus transmission may be affected by HVAC design features in the built environment.

**Methods:** Ovid MEDLINE and Compendex were searched from inception to January 2021. Two reviewers independently screened titles and abstracts and full text of potentially relevant reviews, using a priori inclusion criteria. Inclusion criteria were systematic reviews examining effects of HVAC design features on virus transmission. Two reviewers independently assessed methodological quality using AMSTAR2.

**Results:** Searching identified 361 citations, 45 were potentially relevant, and 7 were included. Reviews were published between 2007 and 2021, and included 47 virus studies. Two earlier reviews (2007, 2016) of 21 studies found sufficient evidence that mechanical ventilation (airflow patterns, ventilation rates) plays a role in airborne transmission; however, both found insufficient evidence to quantify minimum mechanical ventilation requirements. One review (2017) of 9 studies examining humidity and indoor air quality found that influenza virus survival was lowest between 40% and 80% relative humidity; authors noted that ventilation rates were a confounding variable. Two reviews (2021) examined mitigation strategies for coronavirus transmission, finding that transmission decreased with increasing temperature and relative humidity. One review (2020) identified 14 studies examining coronavirus transmission in air conditioning systems, finding HVAC systems played a role in virus spread during previous coronavirus outbreaks. One review (2020) examined virus transmission interventions on public ground transportation, finding ventilation and filtration to be effective.

**Conclusions:** Seven reviews synthesizing 47 studies demonstrate a role for HVAC in mitigating airborne virus transmission. Ventilation, humidity, temperature, and filtration can play a role in viability and transmission of viruses, including coronaviruses. Recommendations for minimum standards were not possible due to few studies investigating a given HVAC parameter. This overview examining HVAC design features and their effects on airborne transmission of viruses serves as a starting point for future systematic reviews and identifying priorities for primary research.

## Introduction

The 2019 novel coronavirus or severe acute respiratory syndrome coronavirus 2 (SARS-CoV-2) outbreak, first detected in Wuhan, China, was characterized as a pandemic by the World Health Organization (WHO) in March 2020 [1]. Almost two years later (early-February 2022), the WHO reported over 383 million cases of the disease (COVID-19) caused by the virus (SARS-CoV-2) with over 5.6 million deaths worldwide [2]. Early in the pandemic there were conflicting views and debate about routes of transmission [3–6]. Several recent reviews of the scientific literature have identified evidence indicating airborne transmission, which could be particularly problematic in confined and/or crowded indoor spaces [7–9]. Public health recommendations acknowledge airborne transmission as important with advice to maximize ventilation, ensure proper maintenance and functioning of heating, ventilation and air conditioning (HVAC) systems, and increase use of fresh air where possible [10].

Airborne transmission occurs as a result of bioaerosols (biological particles suspended in air) staying aloft longer due to their small size and therefore travelling further due to air currents [3]. Several possible mechanisms of airborne coronavirus transmission exist including: 1) bioaerosol generation by infectious persons with coughing, sneezing, breathing and talking, which remain airborne for a period of hours to days; 2) short- to long-range transport through (HVAC systems and subsequent inhalation of bioaerosols by other people; and 3) airborne transport of bioaerosols to surfaces (or contamination of surfaces by physical contact), followed by resuspension, inhalation or contact with surfaces [11, 12].

Previous research demonstrated that infectious airborne bioaerosols spread to other spaces via HVAC systems [12, 13]. Multiple features within HVAC systems may influence transmission, including ventilation (e.g., ventilation rate, air changes per hour (ACH), airflow pattern, pressurization), filtration (e.g., filter minimum efficiency reporting value (MERV) rating, age, extent of use), ultraviolet (UV) radiation (e.g., UV power, UV dose) and humidity [12]. Understanding the influences of HVAC systems on airborne transmission in the built environment is critical for building scientists to develop effective engineering control strategies to protect occupant health and well-being and to affect timely public health policies. Previous systematic reviews provide a starting point for understanding what is already known from the scientific literature about HVAC systems and airborne transmission of viruses. A comprehensive synthesis of previous systematic reviews can also help identify knowledge gaps helping to guide and prioritize future primary research. Therefore, we conducted an overview of reviews to identify and synthesize previous systematic reviews on this topic.

## Methods

Standards recommended by the international Cochrane organization for the conduct of an overview of reviews [14] were followed. The research question guiding this work was: what is the current synthesized evidence about the effects of heating, ventilation and air conditioning (HVAC) design features on virus transmission?

### Search Strategy

A research librarian (GMT) conducted searches in Ovid MEDLINE and Compendex from inception to June 2020, using concepts related to viruses, transmission and HVAC. The search was updated in January 2021. Search strategies appear in Multimedia Appendix 1. The unfiltered search strategies were peer-reviewed by two librarians (TL, AH) and the filter for systematic reviews in Ovid MEDLINE was provided by a third librarian (LD). The unfiltered search strategies were part of a larger systematic review project which is registered [15] and its protocol is publicly available [16]. Reference lists of included reviews were screened to identify any other relevant reviews. Conference abstracts and preprints retrieved through the searches were screened to determine whether a full peer-reviewed manuscript was published. References were managed in EndNote; duplicate records were removed prior to screening.

### Study Selection

Two reviewers (GMT, LH) independently screened the titles and abstracts of all citations retrieved from the electronic searches and other sources. Studies were classified as yes, no or maybe. The first stage of screening was completed in Covidence. We retrieved the full text of all studies classified as yes or maybe. The same reviewers independently applied inclusion/exclusion criteria (Multimedia Appendix 2) to each full text document and classified studies as include or exclude. Discrepancies were resolved through discussion between the two reviewers. Reasons for excluding studies at the full text stage were documented (Multimedia Appendix 3).

### Inclusion and Exclusion Criteria

Inclusion and exclusion criteria are detailed in Multimedia Appendix 2. We planned to include systematic reviews published in English that searched for and included primary research studies examining the effects of HVAC design features on transmission of viruses. The HVAC features of interest were: mechanical ventilation (ventilation rate, air change, air exchange, airflow); filtration (air filtration, filter type, minimum efficiency reporting value (MERV) rating, filter age and/or use, pressure drop, holding capacity, replacement, change frequency); ultraviolet germicidal irradiation, UVGI (power, dose, uniformity of dose, flow rate, bioaerosol inactivation efficiency, location); humidity or relative humidity. Inclusion was staged in two ways. Our primary interest was viruses and we excluded those reviews that were not specific to virus. We were initially interested in systematic reviews defined by the international Cochrane organization as reviews that use a pre-defined, systematic approach and follow standard approaches to search the literature, select studies for inclusion, assess methodological quality of included studies, extract and synthesize/analyze data from the included studies. As we found few systematic reviews meeting these criteria, we included review articles that satisfied specific requirements for methodological approach and objective. For methodological approach requirements, the authors had to search two or more databases, describe inclusion and exclusion criteria, and describe a process for study selection. For objective requirements, the objective of the review had to be related to an HVAC design feature, including ventilation, filtration, ultraviolet radiation, or humidity.

### Quality Assessment

Methodological quality of included reviews was assessed using AMSTAR2 [17]. AMSTAR2 is a valid and reliable tool containing 16 items about the methodological conduct of a systematic review [18]. Two authors (GMT, LH) independently assessed the included reviews. Discrepancies were resolved through discussion.

### Data Extraction

The following information was extracted from each review: citation information (e.g., authors, year of publication, country of corresponding author); objectives; search strategy; inclusion/exclusion criteria; settings; population characteristics (as applicable); agent studied (e.g., type of virus, bioaerosol); HVAC design features studied; number and characteristics of studies relevant to this overview’s research question; results (as reported by review authors) and review authors’ conclusions relevant to this overview’s research question. Our primary outcome was quantitative measures of the association between HVAC design features and virus transmission; however, we extracted any results reported by the review authors that were relevant to our research question. One reviewer (LH) extracted data using a pre-defined form. A second reviewer (EK) verified data. Discrepancies were resolved through discussion and referring to the relevant publication.

### Data Analysis

We anticipated that the included reviews would not have conducted meta-analyses. We planned to present results in tabular and narrative form. Tables were created describing the reviews, their results (including any quantitative data of associations between HVAC features and virus transmission or proxy outcomes) and conclusions, and their methodological quality. A narrative summary of the findings of each review is provided. We only summarize review findings that were relevant to our research question; for example, if the review included studies of ventilation, humidity, etc. in the outdoor and indoor environment, we only report on studies specific to the indoor (built) environment.

## Results

The search retrieved 361 citations, of which 45 were considered potentially relevant and 7 met the inclusion criteria (Figure 1). Tables 1 and 2 provide summaries of the included reviews. The reviews varied somewhat in their objectives (e.g., investigate mechanical ventilation, ventilation rates, airflow patterns, effects of humidity, stability of bioaerosols containing coronaviruses), agents (e.g., coronaviruses, influenza viruses), and settings (e.g., built environment, healthcare settings, public ground transportation). Reviews were published between 2007 and 2021 (median year 2020) and included a total of 47 unique virus studies published between 1961 and 2020 (median year 2005) that were relevant to our research question (median 4 studies per review including shared references, Table 3, Multimedia Appendix 4).

**Figure 1.**
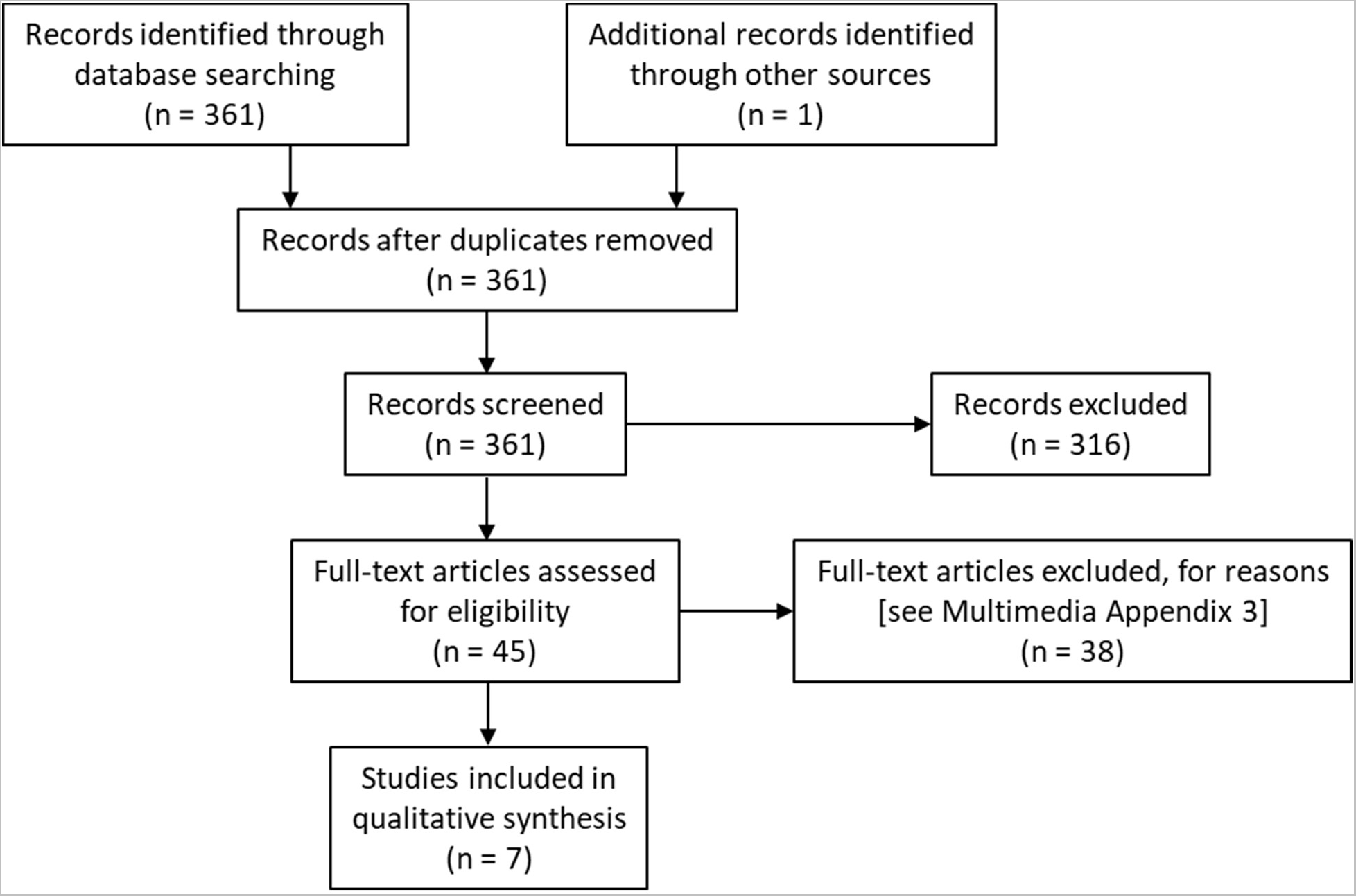
Flow of studies through the selection process.

**Table 1.** Summary of Characteristics of Relevant Reviews.

| Author year; Country<br>Agent; Setting | Purpose / objectives | Search: databases,<br>years | Inclusion criteria | Exclusion Criteria | Study designs |
| --- | --- | --- | --- | --- | --- |
| Li 2007 [13]<br>China<br>Airborne infectious<br>diseases<br>Multiple built<br>environments | “1) Is there sufficient evidence to support that the ventilation rate and/or the airflow pattern are contributing cause(s) for the spread of airborne infectious diseases?<br>2) If so, is there good evidence/ data to support the specification and quantification of minimum ventilation requirements to minimize the transmission of airborne infectious diseases in different settings (nosocomial or otherwise)?” (p.4) | MEDLINE, ISI Web of Knowledge, and ScienceDirect (1960 to March 2005); reviewed references of retrieved articles | “Relevance of the article to the two key research questions” (p.4); “research techniques employed must have been scientifically robust, repeatable and reliable” (p.4); “original articles in English” (p.4) | Conference papers and abstracts; “descriptive articles without an explicit detailed analytic component” (p.4); work before 1960 | Epidemiologic (+/- detailed ventilation studies), case control, cohort, intervention, questionnaire, animal, mathematical modelling |
| Luongo 2016 [19]<br>USA<br>Infectious agents<br>Buildings | To review epidemiologic studies examining the association between ventilation (at least one HVAC parameter) and airborne transmission of infectious agents in buildings, and “to assess the quality and quantity of available data and to identify research needs” (p.666) | Science Direct, Web of Knowledge, MEDLINE/PubMed, Engineering Village, and Google Scholar (search dates not reported) | “Specifically used an epidemiologic study design and that described or measured some HVAC parameter within the context of the hypothesized associations.” (p.666) | Modelling studies | “Epidemiologic studies investigating the association of at least one HVAC-related parameter with an infectious disease-related outcome in buildings (almost all studies reported ventilation rates or CO<sub>2</sub>).” (p.667) |
| Derby 2017 [20]<br>US<br>Multiple infectious<br>agents<br>Laboratory and multiple<br>built environments | “To conduct a broad survey of post-1985 literature regarding the effects of low humidity on comfort, health, and IEQ [indoor environmental quality]” and “to identify existing knowledge and knowledge gaps, as well as confounding variables” (p.30) | Engineering Index (Compendex), Web of Science, Google Scholar; citation search of key papers in Scopus and Google Scholar (search dates not reported); citation checking of relevant review papers | Controlled studies that focus on healthy, human subjects in residences and workplaces with at least one data point where relative humidity 40%, and provide new data and report temperature | Publication post-1985 (unless papers present unique data not previously reviewed); review papers not analyzed in depth | Experimental (laboratory testing studies), transmission studies with animal models, modelling studies, epidemiological studies |
| Chirico 2020 [21] | “to evaluate the COVID-19 risk associated with the presence of air-conditioning systems” (p.2) | PubMed/ MEDLINE, PubMed Central, Google Scholar, | Original studies (observational and experimental) of humans in | Narrative reviews, opinions, commentaries; | Observational and experimental studies (including modelling |
| Italy (corresponding author)<br>SARS-CoV-1/ MERS-CoV/<br>SARS-CoV-2<br>Indoor environments |  | medRxiv (July 11, 2020); cross-referencing | indoor environments, exposed to air-conditioning systems, with respiratory infection outbreaks caused by SARS-CoV-1, MERS-CoV, or SARS CoV-2; English; no time limit | experimental studies on airborne transmission of coronaviruses not associated with outbreaks | and CFD simulation studies) |
| Zhen 2020 [23]<br>South Africa (corresponding author)<br>Viruses such as influenza, SARS-CoV or MERS-CoV<br>Public ground transportation | “to assess the abilities of different interventions to decrease the incidence of droplet-based infections among people using public ground transport” (p.478) | MEDLINE (PubMed), CENTRAL (Cochrane Library), Web of Science (Clarivate Analytics); reference lists of relevant reviews; WHO’s database ‘Global research on coronavirus disease (COVID-19)’ | Interventions (e.g., PPE, etc) and relationship to infections from viruses (e.g., influenza, SARS-CoV or MERS-CoV) in “humans using public transportation (taxis, buses, trains and subways)” (p.479); published between 2000 and 2020 in English. | “participants/context of the intervention were healthcare workers in healthcare facilities” (p.479) | Systematic reviews, clinical trials, comparative observational studies, modelling studies<br>(due to limited relevant research, authors discuss international and national guidance documents) |
| da Silva 2021 [32]<br>Portugal<br>SARS-CoV, MERS-CoV, SARS-CoV-2<br>Indoor and outdoor environments | To discuss “the viability/stability of aerosols containing SARS-CoV and MERS-CoV viruses...to provide information on potential mitigation strategies for SARS-CoV-2 airborne transmission” (p.2) | PubMed/MEDLINE, Web of Science and Scopus; references of studies were screened | Studies published since 2002 (emergence of SARS-CoV); virus studied was SARS-CoV, MERS-CoV or SARS-CoV-2; viability of the virus sampled from air was assessed; no language limits | n/a | Real-world sampling and laboratory studies |
| Noorimotlagh 2021 [36]<br>Iran<br>Human coronaviruses (HCoV)<br>Laboratory experimental set-ups | “to collect all available studies concerning inactivation methods, environmental survival, and control and prevention strategies” (p.1) | Scopus, ISI Web Science, Google Scholar, PubMed (MEDLINE), World Health Organization, American Centers for Disease Control and Prevention; 1990-2020 | Original studies; published in English; available electronically (online); focus on disinfections, environmental survival, and control and prevention strategies of HCoV | Review articles, book review, guidelines, book chapters, duplicate articles, short communications, conference documents, oral presentation, comments | Original research (study designs not described, appear mostly experimental laboratory-based studies) |

CFD = computational fluid dynamics; HCoV = human coronaviruses; HVAC = heating, ventilation, and air conditioning; ISI = Institute for Scientific Information; n/a = not applicable; PPE = personal protective equipment

**Table 2.** Summary of Results and Conclusions from Relevant Reviews.

| Author year | Results | Conclusions |
| --- | --- | --- |
| Li 2007 [13] | Based on multi-disciplinary consensus panel: “of the 40 studies, 18 were considered as non-conclusive or not meeting evidentiary threshold to support a direct contributory role of ventilation rate/airflow pattern to the airborne spread of infectious agents, 12 were partly conclusive or met threshold somewhat, 10 were deemed clearly conclusive supporting a direct contribution.” (p.5) | “There is insufficient data to specify and quantify the minimum ventilation requirements in hospitals, schools, offices, homes and isolation rooms in relation to spread of infectious diseases via the airborne route.” (p.2)<br>“There is strong and sufficient evidence to demonstrate the association between ventilation, air movements in buildings and the transmission/spread of infectious diseases such as measles, tuberculosis, chickenpox, influenza, smallpox and SARS.” (p.15) |
| Luongo 2016 [19] | 13 studies (1988-2013) of which 11 were observational and 2 were intervention studies. Building-related factors (e.g., ventilation rates) were associated with increased measures of illness in 11 studies; one study showed no association and one was inconclusive. | “Studies to date show an association between increased infectious illness and decreased ventilation rate, however, there are insufficient data to quantify how mechanical ventilation may affect the airborne transmission of infectious agents.” (p.666)<br>“The weight of the data implies that HVAC system factors in buildings have a role in APT; however, more studies need to be completed, with the eventual goal of a meta-analysis to integrate results.” (p.676) |
| Derby 2017 [20] | About 70 articles included overall; 9 papers examined effects of humidity on viability or transmission of airborne viruses.<br>4 studies showed decreased virus viability at mid-range (~50%) RH.<br>5 studies showed “a canonical dip between 40 and 80% RH” (p.39).<br>3 studies suggested greater transmission at lower humidity (e.g., 20-35% vs 50% RH).<br>One study showed importance of ventilation rates in removing airborne viruses, especially in smaller droplets. | Influenza virus survival dips between 40 and 80% RH.<br>“Lower humidity increased virus survival for influenza.” (p.30)<br>Survival declines with increased length of exposure.<br>Across many low humidity studies, ventilation rates and exposure times were noted as confounding variables.” (p.30) |
| Chirico 2020 [21] | 14 studies of outbreaks associated with air-conditioning systems, all in Far East (Asian countries); 6/7 studies on SARS-CoV-1 indirectly proved role of HVAC; 1 study of MERS showed contamination of HVAC; 4/6 studies on SARS-CoV-2 diffusion of virus through HVAC suspected or supported by computer simulation | Evidence of HVAC systems facilitating spread of coronaviruses in previous outbreaks in Asian (Far East) countries.<br>Evidence for SARS-CoV-2 is limited and does not provide sufficient evidence that SARS-CoV-2 can be transmitted by HVAC systems. Generalization of results to other regions limited due to technological differences in HVAC systems. |
| Zhen 2020 [23] | 4 studies included; 1 SR showed use of public transportation increased risk of influenza transmission; 1 case-control study did not show increased risk of influenza diagnosis with use of public transport; 2 modelling studies showed airborne infection on trains can be reduced with facemasks, adequate ventilation, and filtration in cases of where non-recirculated air not possible | “filtering air being circulated within the bus can reduce airborne transmission of influenza between passengers, and improving ventilation on a train can decrease the risk of influenza infection” (p.482)<br>Public transport increases risk of transmission of influenza; risk increases with trip duration and proximity to an infected individual; modelling studies suggest that adequate ventilation could reduce transmission risk. |
| da Silva 2021 [32] | 11 studies included; 8 air sampling; 3 laboratory-based experimental studies : 1 MERS-CoV study showed decreased stability at 70% RH compared with 40% RH at 20 °C; 1 MERS-CoV study found high robustness and strong capability to survive (63.5% of viruses remaining infectious 60 min after aerosolization) at 25 °C and 79% RH; 1 SARS-CoV-2 study showed aerosol survival time of 3 h | “Temperatures ranging from 20 °C to 25 °C and relative humidity ranging from 40% to 50% were reported to have a protective effect on viral viability for airborne SARS-CoV and MERS-CoV.” (p.1)<br>“higher temperatures and high relative humidity can have an effect on SARS-CoV-2 viability in the environment as reported in previous studies” (p.8)<br>(conclusions relate to both indoor and outdoor environments) |
| Noorimotlagh 2021 [36] | 42 studies (20 of inactivation/ disinfection methods; 12 of environmental survival; 10 of prevention and control strategies); 1 study of Phi6 showed highest virus survival at RH >85% and <60% with significant decrease ~60-85%, At fixed RH of 75%, infectivity decreased two orders of magnitude between 19°C and 25°C; 1 study where aerosolized MERS-CoV data reported in da Silva 2021 [32]; 1 study of MERS-CoV found robustness and strong capability to survive at 25 °C and 79% RH; 1 study showed aerosol survival time for SARS-CoV-2 of 3 h at 40% RH and 21-23°C, and stability of SARS-CoV-2 similar to SARS-CoV-1 | “temperature and relative humidity are important factors in the survival of SARS-CoV-2” (p.1)<br>“disease transmission via droplets is inhibited by increasing both temperature and RH in buildings” (p.11)<br>SARS-CoV-2 can survive in aerosols for approximately 3 hours.<br>Recommend “proper ventilation of the buildings in time of aerosol generating” (p.13) (though do not review studies of ventilation) |

APT = airborne pathogen transmission; HVAC = heating, ventilation, and air conditioning; RH = relative humidity

**Table 3.** Relevant studies from included reviews that are pertinent to overview research question.

| Citation | Li<br>2007<br>[13] | Luongo<br>2016<br>[19] | Derby<br>2017<br>[20] | Chirico<br>2020<br>[21] | Zhen<br>2020<br>[23] | da Silva<br>2021<br>[32] | Noorimotlagh<br>2021<br>[36] | Topic(s) |
| --- | --- | --- | --- | --- | --- | --- | --- | --- |
| Akers T. *Appl Microbiol* 1966;14:361-4.[49] |  |  | X |  |  |  |  | Humidity |
| Bloch AB. *Pediatr* 1985;75:676-83.[50] | X |  |  |  |  |  |  | Ventilation (airflow) |
| Browne A. *J Travel Med* 2016;18:1-7.[24] |  |  |  |  | X |  |  | Ventilation (ventilation rate) |
| Castilla J. *Influenza Other Respir Viruses* 2013;7:177-83.[52] |  |  |  |  | X |  |  | Virus survival/detection |
| Chen C. *J R Soc Interface* 2011;8:699-710.[53] |  |  |  | X |  |  |  | Ventilation (airflow) |
| de la Noue AC. *Appl Environ Microbiol* 2014;80:7196-205.[54] |  |  | X |  |  |  |  | Humidity |
| Drinka PJ. *J Am Geriatr Soc* 1996;44:910-13.[41]<sup>1,4</sup> | X | X |  |  |  |  |  | Ventilation (airflow); Filtration |
| Drinka PJ. *Infect Control Hosp Epidemiol* 2002;23:600-3.[55]<sup>1</sup> | X |  |  |  |  |  |  | Ventilation (airflow) |
| Drinka PJ. *J Am Geriatr Soc* 2004;52:847-48.[42]<sup>1,4</sup> | X | X |  |  |  |  |  | Ventilation (ventilation rate) |
| Furuya H. *Environ Health Prev Med* 2007;12:78-83.[56] |  |  |  |  | X |  |  | Ventilation (ventilation rate) |
| Gustafson TL. *Pediatr* 1982;70:550-6.[57] | X |  |  |  |  |  |  | Ventilation (airflow) |
| Harper GJ. *J Hyg* 1961;59:479-86.[58] |  |  | X |  |  |  |  | Humidity |
| Hemmes JH. *Antonie Van Leeuwenhoek* 1962;28:221-33.[59] |  |  | X |  |  |  |  | Humidity |
| Kim SH. *Clin Infect Dis* 2016;63:363-69.[22] |  |  |  | X |  | X |  | Ventilation (airflow, ventilation rate) |
| Le DH. *Emerg Infect Dis* 2004;10:265-68.[60] | X |  |  |  |  |  |  | Ventilation (airflow) |
| Leclair JM. *N Engl J Med* 1980;302:450-53.[61] | X |  |  |  |  |  |  | Ventilation |
| Lee N. *N Engl J Med* 2003;348:1986-94.[62] |  |  |  | X |  |  |  | Virus survival/detection |
| Li Y. *Indoor Air* 2005;15:83-95.[39]<sup>2</sup> | X |  |  | X |  |  |  | Ventilation (airflow, ventilation rate) |
| Li Y. *Indoor Air* 2005;15:96-111.[45] | X |  |  | X |  |  |  | Ventilation (airflow) |
| Li Y. *medRxiv* 2020.[63] |  |  |  | X |  |  |  | Ventilation (ventilation rate) |
| Lowen AC. *PLoS Pathog* 2007;3:e151.[64] |  |  | X |  |  |  |  | Humidity |
| Lowen AC. *J Virol* 2014;88:7692-5.[65] |  |  | X |  |  |  |  | Humidity |
| Lu J. *Emerg Infect Dis* 2020;26:1628-31.[66] |  |  |  | X |  |  |  | Ventilation (ventilation rate) |
| Mizumoto K. *Infect Dis Model* 2020;5:264–70.[67] |  |  |  | X |  |  |  | Ventilation (airflow) |
| Moser MR. *Am J Epidemiol* 1979;110:1-6.[68] | X |  |  |  |  |  |  | Ventilation (ventilation rate) |
| Myatt TA. *Am J Respir Crit Care Med* 2004;169:1187-90.[43] | X | X |  |  |  |  |  | Ventilation (ventilation rates) |
| Noti JD. *PloS One* 2013;8:e57485.[69] |  |  | X |  |  |  |  | Humidity |
| Olsen SJ. *N Engl J Med* 2003;349:2416-22.[70] | X |  |  |  |  |  |  | Ventilation (ventilation rate) |
| Prussin AJ. *Appl Environ Microbiol* 2018;84:e00551–18.[71] |  |  |  |  |  |  | X | Humidity |
| Pyankov OV. *J Aerosol Sci* 2018;115:158-63.[34] |  |  |  |  |  | X | X | Humidity |
| Qian H. *medRxiv* 2020.[70] |  |  |  | X |  |  |  | Ventilation (ventilation rates) |
| Remington PL. *JAMA* 1985;253:1574-7.[71] | X |  |  |  |  |  |  | Ventilation (ventilation rate) |
| Riley RL. *Am J Epidemiol* 1978;107:421-32.[72]<sup>3</sup> | X |  |  |  |  |  |  | Ventilation (airflow) |
| Riley RL. *Bull Eur Physiopathol Respir* 1979;15:699-705.[73]<sup>3</sup> | X |  |  |  |  |  |  | Ventilation (airflow) |
| Schulman JL. *Nature* 1962;195: 1129-30.[74] | X |  |  |  |  |  |  | Humidity |
| van Doremalen N. *Euro Surveill* 2013;18:1-4.[33] |  |  |  |  |  | X | X | Humidity |
| van Doremalen N. *N Engl J Med* 2020;382:1564-67.[35] |  |  |  |  |  | X | X | Virus survival/detection |
| Wehrle PF. *Bull World Health Organ* 1970;43:669-79.[75] | X |  |  |  |  |  |  | Humidity |
| Wong TW. *Emerg Infect Dis* 2004;10:269-76.[38]<sup>2</sup> | X | X |  | X |  |  |  | Ventilation (airflow); Humidity |
| Xu P. *medRxiv* 2020.[76] |  |  |  | X |  |  |  | Ventilation |
| Yang W. *PloS One* 2011;6:e21481.[77] |  |  | X |  |  |  |  | Humidity |
| Yang W. *PloS One* 2012;7:e46789.[78] |  |  | X |  |  |  |  | Humidity |
| Yu IT. *N Engl J Med* 2004;350:1731-39.[44] | X |  |  | X |  |  |  | Ventilation (airflow) |
| Yu ITS. *Clin Infect Dis* 2005;40:1237-43.[40]<sup>2</sup> | X |  |  | X |  |  |  | Ventilation (airflow) |
| Zhang I. *Emerg Infect Dis* 2013;19:1403-10.[79] |  |  |  | X |  |  |  | Ventilation (airflow) |
| Zhu S. *Build Environ* 2012;47(1):67-75.[80] |  |  |  |  | X |  |  | Ventilation (airflow) |
| Zitter JN. *JAMA* 2002;288:483-86.[81] | X |  |  |  |  |  |  | Ventilation (airflow) |
| Total number of studies relevant to this overview per included review | 21 | 4 | 9 | 14 | 4 | 4 | 4 |  |

1. Li et al [13] evaluates Drinka et al [41], Drinka et al [55], and Drinka et al [42] as one. 2. Li et al [13] evaluates Li et al [39], Wong et al [38], and Yu et al [40] as one. 3. Li et al [13] evaluates Riley et al [72] and Riley et al [73] as one. 4. Luongo et al [19] evaluates Drinka et al [41] and Drinka et al [42] as one.

Li et al [13] examined the role of ventilation (specifically ventilation rates and airflow patterns) in airborne transmission of infectious agents in indoor settings. The authors included 40 English language studies overall, with 16 specific to viruses reported between 1962 and 2005 (median year 1985/1996). Three of the 16 studies included multiple papers (Table 3) which increased the total count to 21. Sixteen studies were epidemiologic, four involved other observational designs, and one was experimental. Three studies had limited and four had no investigation of ventilation rates or airflow. Studies involved a variety of settings: hospital, hospital ward, or health clinic (n=9); aircraft (n=3); nursing home (n=3); school (n=2); high-rise apartments (n=2); offices (n=1); and animal cage (n=1). Viral agents included SARS-CoV-1 (n=7), influenza (n=5), measles (n=4), chicken pox (n=2), rhinovirus (n=1), common cold (n=1), and smallpox (n=1). Overall quality was assessed as good for twelve, average for five, and unsatisfactory for four studies. The researchers convened a panel of experts in medicine, public health, and engineering. They used a modified Delphi approach with final consensus meeting to rate the ‘evidentiary threshold’ to support their hypothesis, i.e., direct contribution of ventilation to airborne transmission. Among the virus studies, eight were rated as conclusive, while eight were partly conclusive, and five were non-conclusive. Among the eight conclusive studies, two examined ventilation rates showing higher rates of infection for influenza with lower ventilation rates and six demonstrated an association between airflow patterns and transmission of measles (pediatric office suite), chicken pox (hospital), smallpox (hospital), and SARS-CoV-1 (hospital). In all studies the bioaerosols travelled a “considerable distance” which the reviewers note “seemed to be related to building design” [13,p.12] (e.g., placement of heating radiator, room pressure, functional status of return air outlet). None of the virus studies provided data to support “specification and quantification of the minimum ventilation requirements” [13,p.3].

Luongo et al [19] examined evidence from epidemiological studies for the association of mechanical ventilation (at least one HVAC parameter) with airborne transmission of infectious agents in buildings. While the authors included 13 English-language studies, 3 were specific to viruses; all studies were observational and were reported between 1996 and 2011 (median year 2004). One study had two papers which increased the total count to 4 (Table 3). All four virus studies were also included in Li et al [13]. Settings included nursing home (n=2), office buildings (n=1), and hospitals (n=1). Viruses represented in the studies included influenza (n=2), SARS-CoV-1 (n=1), and rhinovirus (n=1). Review authors did not assess methodological quality but provided a narrative commentary of the strengths and limitations of each study. Two of the four studies found an association between virus incidence rates, self-reported incidence rates, and risk of exposure with HVAC design features. In a retrospective cohort study of a SARS-CoV-1 outbreak in a hospital, authors measured ventilation rates and found that “proximity to index patient associated with transmission” [19,p.668]. Authors of the second study blindly adjusted outdoor air supply dampers in three office buildings and found a significant positive association between average CO_2_ greater than 100 ppm above background and frequency of rhinovirus detection in air filters. The third study found lower incidence of influenza in newer nursing homes that had 100% outside air delivery (compared to older homes with 30-70% recirculated air) and filtered room supply (compared with no filtration) during one season; however, data collected over five subsequent years, reported in the fourth study, found no clear association. None of the studies quantified minimum ventilation requirements.

Derby et al [20] conducted a literature review to assess the effects of low humidity (less than or equal to 40% relative humidity or RH) on comfort, health, and indoor environmental quality. While the review included around 70 papers, 9 papers examined the effects of humidity on the viability or transmission of airborne viruses. Seven studies were experimental (involving laboratory testing), one paper was a re-analysis of data from one of the experimental studies, and one study involved modelling. Most studies focused on influenza, with one study each examining Columbia SK viruses, murine norovirus, and multiple viruses (influenza, vaccinia, Venezuelan equine encephalomyelitis and poliomyelitis). Most studies examined a wide range of relative humidity from approximately 5-25% RH at the lower range to 75-100% RH at the upper range. The absolute humidity was approximately 25 g/m^3^ or less in all studies except one (which ranged from 25 to 125 g/m^3^). Review authors did not assess the methodological quality of the included studies. In terms of virus viability, four studies showed a reduction at mid-range RH (i.e., approximately 50% RH). Review authors further noted that five studies showed that “virus survival exhibited a canonical dip between 40 and 80% RH” [20,p.39], and that in almost all cases decline in survival was correlated with increased length of exposure. Three studies examined influenza transmission. One study showed reduced transmission among guinea pigs at 50% versus 20 to 35% RH; however, the same pattern was not found when the researchers analyzed the data based on absolute humidity. A second study examined transmission via coughing using manikins and found five times more infectious virus at 7-23% RH versus greater than 43% RH. A modelling study of influenza virus transmission via coughing showed that the infectious virus concentration was 2.4 times more at 10% RH versus 90% RH after 10 minutes and the ratio increased over time. They also demonstrated that the effect of humidity related to particle size: settling of larger particles and inactivation of smaller particles (<5 µm) with greater humidity. They concluded that inactivation resulting from high relative humidity coupled with ventilation is important to remove smaller particles.

Chirico et al [21] conducted a rapid review (streamlined systematic review methods) to examine the potential role of air conditioning (HVAC) systems in “outbreaks of coronaviruses (SARS-CoV-1, MERS-CoV, SARS-CoV-2) in indoor environments” [21,p.2]. The authors identified 14 studies published between 2003 and 2020 (11 peer-reviewed, 3 pre-prints all concerning SARS-CoV-2); studies investigated outbreaks in Hong Kong (n=7), South Korea (n=1), Japan (n=3), and China (n=3). Seven studies examined two outbreaks associated with SARS-CoV-1: five studies examined outbreaks (different areas or groups of individuals) within the same hospital and 2 studies investigated an outbreak in the same private high-rise housing estate. Five of the SARS-CoV-1 studies from Chirico et al [21] are shared references with Li et al [13] (Table 3). Review authors indicated that six of the seven studies indirectly demonstrated a role of the HVAC system (through epidemiological data, spatiotemporal patterns of infection, or modelling). One study investigated an outbreak of MERS-CoV in a hospital setting and demonstrated contamination of the HVAC system through environmental sampling [22]. Six studies investigated outbreaks of SARS-CoV-2: one study examined 318 outbreaks in 120 cities in China including community and workplace settings; three studies examined an outbreak on a ship in Japan; and two studies examined the same outbreak in a restaurant. Three observational studies suspected a role of the HVAC system, two studies (both of ship outbreak) did not find evidence of a role of HVAC based on the spatiotemporal distribution of cases, and one study (of restaurant outbreak) showed support of the role of HVAC by computer simulation. Review authors indicated that they were not able to appropriately evaluate the quality of the included studies. Review authors concluded that there is sufficient evidence from SARS-CoV-1 and MERS-CoV demonstrating a role of HVAC in airborne transmission of the viruses; however, there was not sufficient evidence that HVAC systems play an important role in the case of SARS-CoV-2. While there is a lack of evidence for SARS-CoV-2, there is not evidence of no role.

Zhen et al [23] conducted a rapid review of “the role of public ground transport in COVID-19 transmission” and “interventions that may reduce transmission” [23,p.478]. The authors searched for studies published since 2000 and identified four relevant studies, published between 2007 and 2016, including a systematic review, a case-control study, and two modelling studies. The systematic review by Browne et al [24] identified 41 studies examining the risk of transmission of Influenza A (H1N1/09) (29 studies), SARS-CoV (5 studies), or MERS-CoV (2 studies) related to sea (6 studies), ground (6 studies), or air (30 studies) transport. Zhen et al [23] summarized results from four quantitative studies included in Browne et al [24], and concluded that “use of public transport increased the risk of influenza transmission” [23,p.481]. Zhen et al [23] identified a multi-centre case-control study that showed a lower probability of Influenza A (H1N1/09) diagnosis with public transport use (metro, bus, tram or local train), and no association with use of train, aeroplane or taxi. The case-control study was assessed as moderate risk of bias by Zhen et al [23]; risk of bias was not reported for the other three studies. Zhen et al [23] also identified two modelling studies, one estimated the reproduction number for influenza infection in a train and the other tested simulations to predict influenza infection probability for four bus ventilation systems. The first modelling study showed that masks could decrease reproduction number resulting in lower risk of disease transmission, with high efficiency particulate air (HEPA) masks more effective than surgical masks. Further, doubling the ventilation rate reduced risk similar to use of HEPA masks, and was considered more feasible and cost-effective. The second modelling study showed that influenza transmission risk can be reduced when the infected passenger is positioned closer to the exhaust opening and with high efficiency filtration in the case where non-recirculated air cannot be provided. Given the limited number of research studies, Zhen et al [23] also identified and discussed national and international guidance documents, e.g., WHO, etc. [25–31]. While general recommendations have been made to reduce risk (e.g., minimize use of public transport, environmental controls, respiratory etiquette, hand hygiene, mask use), there is no indication of the empirical evidence specific to these measures, in particular mechanical ventilation.

Da Silva et al [32] conducted a systematic review to discuss “the viability/stability of aerosols containing SARS-CoV and MERS-CoV viruses” with an intent “to provide information on potential mitigation strategies for SARS-CoV-2 airborne transmission” [32,p.2]. The review authors identified 11 studies: eight studies examined the viability of coronaviruses in air samples but review authors did not describe the relationship with HVAC features, including one MERS-CoV study [22] which was described above by Chirico et al [21]. Three studies were laboratory-based experimental studies of coronaviruses. In one MERS-CoV study the virus was aerosolized at 20 °C with 40% or 70% RH, showing decreased stability at 70% RH compared with 40% RH [33]. The other MERS-CoV study examined virus inactivation under two conditions [34]: common office environment (25 °C and 79% RH) and Middle Eastern region climate (38 °C and 24% RH). In the simulated office environment, “the virus demonstrated high robustness and strong capability to survive with about 63.5% of viruses remaining infectious 60 min after aerosolisation. Virus decay was much stronger for hot and dry air scenario with only 4.7% survival over 60 min procedure.” [32,p.5-6]. One study showed aerosol survival time for SARS-CoV-2 of 3 h [35]. Review authors did not assess the methodological quality of included studies; however, they commented on some limitations. The review authors concluded that “higher temperatures and high relative humidity can have an effect on SARS-CoV-2 viability in the environment as reported in previous studies to this date” [32,p.8]. However, their conclusions are based on studies of both indoor and outdoor environments.

Noorimotlagh et al [36] performed a systematic review of SARS-CoV-2 literature “to collect all available studies concerning inactivation methods, environmental survival, and control and prevention strategies” [36,p.1]. While 42 studies were identified, four provided information on temperature and humidity in the built environment, investigating, MERS-CoV (n=2), SARS-CoV-1 and SARS-CoV-2 (n=1), and Phi6 (n=1) which is a bacteriophage used as a surrogate for virus. All were laboratory-based experimental studies. Review authors did not assess or comment on the methodological quality of the studies they included. The aerosolized MERS-CoV data from van Doremalen et al [33] was reported in da Silva et al [32] although not extracted by Noorimotlagh et al [36]. A MERS-CoV study found robustness and strong capability to survive at 25°C and 79% RH [34]; this was a shared reference with da Silva et al [32]. While da Silva et al [32] reported the SARS-CoV-2 survival time from van Doremalen et al [35], Noorimotlagh et al [36] further clarified that the aerosol survival time for SARS-CoV-2 of 3 h was at 40% RH and 21-23°C, and that the stability of SARS-CoV-2 was similar to SARS-CoV-1 [35]. The Phi6 study showed highest virus survival at greater than 85% RH and less than 60% RH with significant decrease between 60% RH and 85% RH [37]. At fixed humidity of 75% RH, infectivity decreased two orders of magnitude between 19°C and 25°C [37]. The review authors concluded that “temperature and relative humidity are important factors in the survival of SARS-CoV-2” [36,p.1] and that “disease transmission via droplets is inhibited by increasing both temperature and RH in buildings” [36,p.11]. One review recommendation was “proper ventilation of the buildings in time of aerosol generating” [36,p.13]; however, studies of ventilation were not reviewed.

The network of the seven included reviews and their 47 references relevant to this overview was created using Palladio (Figure 2). Overall, 12 references were shared between the seven included reviews. However, the network clearly demonstrates that Derby et al [20] and Zhen et al [23] share no references with the five other reviews. In actuality, the 12 shared references were shared between 5 reviews (Figure 2). da Silva et al [32] and Noorimotlagh et al [36] share three references regarding experimental studies of MERS-CoV and SARS-CoV-2 [33–35] (Table 3). Also, da Silva et al [31] shares one reference on MERS-CoV isolation wards [22] with Chirico et al [21]. Three reviews, Li et al [13], Luongo et al [19] and Chirico et al [21], share a reference on SARS-CoV-1 in hospital wards [38]. Li et al [13] and Chirico et al [21] share other related references on SARS-CoV-1 in hospital wards [39, 40]. Similarly, Li et al [13] and Luongo et al [19] share studies on influenza in nursing homes [41, 42] and rhinovirus in offices [43]. Li et al [13] and Chirico et al [21] share two other references regarding SARS-CoV-1 in high-rise apartment complexes [44, 45]. Not only were the 12 shared references only shared between 5 reviews, eight of the shared references were shared with one 2007 review, Li et al [13] and the remaining 4 shared references were shared with one 2021 review, da Silva et al [32] (Figure 2). While 35 of the 47 references were not shared, the 12 shared references were captured by the earliest review (2007 [13]) and one of the latest reviews (2021 [32]).

**Figure 2.**
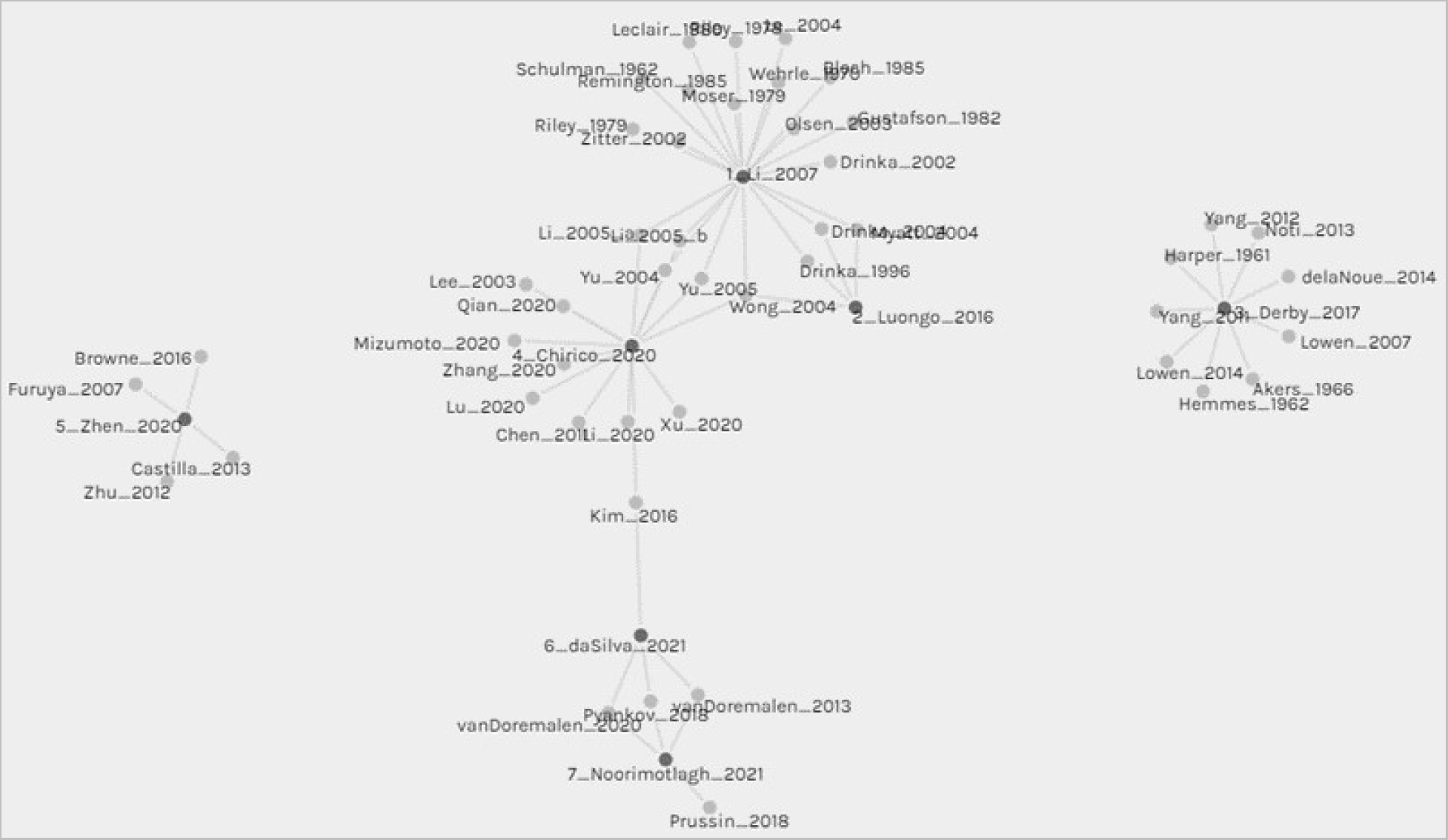
Network representing relevant references (grey circles) from the seven included reviews (black circles): 1_Li_2007; 2_Luongo_2016; 3_Derby_2017; 4_Chirico_2020; 5_Zhen_2020, 6_daSilva_2021; 7_Noorimotlagh_2021. Shared references: 1_Li_2007, 2_Luongo_2016 and 4_Chirico_2020 share Wong_2004 1_Li_2007 and 2_Luongo_2016 share Drinka_1996; Drinka 2004; Myatt 2004 1_Li_2007 and 4_Chirico_2020 share Li_2005_a; Li_2005_b; Yu_2004; Yu_2005 4_Chirico_2020 and 6_daSilva_2021 share Kim_2016 6_daSilva_2021 and 7_Noorimotlagh_2021 share vanDoremalen_2013; Pyankov_2018; vanDoremalen_2020

Table 4 provides the assessments of the methodological quality of the reviews based on AMSTAR2. Three papers described themselves as systematic reviews, two were rapid reviews, one was a broad literature survey, and one was described simply as a review. The majority provided detailed research questions, explained study designs considered for inclusion, used a comprehensive search strategy, described the included studies, discussed heterogeneity of results, and reported potential conflicts of interest. None or few reviews provided an a priori protocol, performed study selection and data extraction in duplicate, provided a list of excluded studies, conducted risk of bias assessments of individual studies, or reported on sources of funding for included studies. None of the reviews conducted a meta-analysis; all provided a narrative synthesis of results/observations across included studies. A previous review [19] spoke about the need for more well-designed studies (including representative sampling, clear and consistent measurement methods and reporting of data) with the goal of meta-analysis to integrate results.

**Table 4.** Methodological Quality of Relevant Reviews based on AMSTAR2.

| AMSTAR2 Question | Li<br>2007<br>[13] | Luongo<br>2016<br>[19] | Derby<br>2017<br>[20] | Chirico<br>2020<br>[21] | Zhen<br>2020<br>[23] | da Silva<br>2021<br>[32] | Noorimotlagh<br>2021<br>[36] |
| --- | --- | --- | --- | --- | --- | --- | --- |
| 1. Did the research questions and inclusion criteria for the review include the components of PICO? | Yes | Yes | Yes | Yes | Yes | Yes | Yes |
| 2. Did the report of the review contain an explicit statement that the review methods were established prior to the conduct of the review and did the report justify any significant deviations from the protocol? | No | No | No | No | No | No | No |
| 3. Did the review authors explain their selection of the study designs for inclusion in the review? | Yes | Yes | Yes | Yes | Yes | No | No |
| 4. Did the review authors use a comprehensive literature search strategy? | Yes | Partial Yes | Partial Yes | Yes | Yes | Yes | Partial Yes |
| 5. Did the review authors perform study selection in duplicate? | Yes | No | No | No | No | Yes | Yes |
| 6. Did the review authors perform data extraction in duplicate? | No | No | No | No | No | No | No |
| 7. Did the review authors provide a list of excluded studies and justify the exclusions? | No | No | No | No | No | No | No |
| 8. Did the review authors describe the included studies in adequate detail? | Yes | Yes | Partial yes | Yes | Yes | Yes | Yes |
| 9. Did the review authors use a satisfactory technique for assessing the risk of bias (RoB) in individual studies that were included in the review? | No | No | No | No | Partial yes | No | No |
| 10. Did the review authors report on the sources of funding for the studies included in the review? | No | No | No | No | No | No | No |
| 11. If meta-analysis was performed did the review authors use appropriate methods for statistical combination of results? | No meta-analysis conducted | No meta-analysis conducted | No meta-analysis conducted | No meta-analysis conducted | No meta-analysis conducted | No meta-analysis conducted | No meta-analysis conducted |
| 12. If meta-analysis was performed, did the review authors assess the potential impact of RoB in individual studies on the results of the meta-analysis or other evidence synthesis? | No meta-analysis conducted | No meta-analysis conducted | No meta-analysis conducted | No meta-analysis conducted | No meta-analysis conducted | No meta-analysis conducted | No meta-analysis conducted |
| 13. Did the review authors account for RoB in individual studies when interpreting/ discussing the results of the review? | Yes | Yes | No | No | Yes | Yes | No |
| 14. Did the review authors provide a satisfactory explanation for, and discussion of, any heterogeneity observed in the results of the review? | Yes | Yes | Yes | Yes | Yes | Yes | No |
| 15. If they performed quantitative synthesis did the review authors carry out an adequate investigation of publication bias (small study bias) and discuss its likely impact on the results of the review? | No meta-analysis conducted | No meta-analysis conducted | No meta-analysis conducted | No meta-analysis conducted | No meta-analysis conducted | No meta-analysis conducted | No meta-analysis conducted |
| 16. Did the review authors report any potential sources of conflict of interest, including any funding they received for conducting the review? | Yes | Yes | Yes | Yes | Yes | Yes | Yes |

## Discussion

### Principal Findings

This comprehensive overview of reviews provides a map of the existing synthesized evidence on the role of HVAC in airborne virus transmission. The earliest review by Li et al [13] published in 2007 found evidence of an association between ventilation rates and airflow patterns in buildings and transmission of viral diseases. Li et al [13] found no studies that provided minimum ventilation requirements to prevent the spread of viral diseases; however, they found one study showing tuberculin conversion was significantly associated with ventilation rates of less than 2 ACH in general patient rooms [44]. Published in 2007 shortly after the 2003 SARS-CoV-1 epidemic, Li et al called for a “multidisciplinary research culture” [13,p.14] to study outbreaks, as well as smaller scale transmission occurrences, to fill the gap with respect to quantifying minimum ventilation standards in both clinical and non-clinical settings. A subsequent review by Luongo et al [19] published almost ten years later in 2016 included a subset of four of the virus studies identified by Li et al [13] with similar conclusions about the possible association between ventilation features (low outdoor air supply, imbalance in supply and exhaust airflow rates) and airborne virus transmission. Luongo et al [19] also pointed out the lack of data to quantify how mechanical ventilation may affect airborne transmission and the need for more well-designed multi-disciplinary epidemiological studies. More recently in response to the current COVID-19 pandemic, Chirico et al [21] examined HVAC systems and their role in airborne transmission of coronaviruses; they concluded there was sufficient evidence demonstrating an association for SARS-CoV-1 and MERS-CoV while there was a lack of evidence for SARS-CoV-2. Derby et al [20] specifically examined the role of humidity in relation to indoor air quality; the evidence they identified was specific to influenza and showed virus survival was lowest between 40% and 80% RH and survival time decreased with length of exposure to humidity. One of the studies from Noorimotlagh et al [36] indicated that aerosolized SARS-CoV-2 can survive for 3 h at 40% RH and 21-23 °C [35]. In another recent review published in 2021, da Silva et al [32] examined mitigation strategies and found two studies demonstrating that coronavirus transmission decreased with increasing both temperature and relative humidity in buildings. A recent review (2020) by Zhen et al [23] examined interventions to reduce virus transmission on public ground transportation; two modelling studies showed ventilation and filtration to be effective.

### Comparison with Prior Work

While there is an extensive body of literature examining HVAC and its role in airborne virus transmission, there is lack of empirical evidence to quantify minimum standards for HVAC design features in the built environment. The previous reviews have discussed this gap, stressed the need for methodologically rigorous epidemiological studies involving multiple disciplines (e.g., engineering, medicine, epidemiology, and public health), and discussed considerations for future research. These include specificity of the virus, its construction and envelope composition, the infectious dose, and the particle size containing the virus. Review authors have called for standardizing experimental conditions, measurements, terminology, and reporting, as well as simulating real world conditions [19,20,32]. An important consideration in designing rigorous studies is controlling for confounding factors. HVAC systems operate in a complex environment; for example, Derby et al [20] noted several confounding variables to be considered when interpreting their findings on humidity and temperature including “variation in air exchange rate, length of organism exposure, variation in the biological structure and routes of entry, variation of pathogen survival on different fomites, and variances in human host response” [20,p.33]. They further noted that the number and complexity of variables to consider “greatly increases the test matrices required” [20,p.39] to build a comprehensive evidence base. Studies have also demonstrated the importance of the positioning of the infected person relative to HVAC features and other occupants, mobility patterns and activities (e.g., type and intensity of respiratory activity) of occupants, time spent within a space, occupancy and occupant density. Despite specification of air flow parameters, the flow of air in occupied spaces is almost always turbulent (versus laminar) such that particles “are constantly mixing and moving in varied ways across a space” making assessments and predictions challenging [47]. Finally, research results need to be interpreted in light of technological differences in HVAC systems around the world [21]. Engineers have developed sophisticated methods (through modelling, computational fluid dynamics, etc.) that allow for isolation of features and control for confounding variables. However, these studies rely on many assumptions that may not hold in real world settings or are specific to an assumed building design or configuration. As well, these studies may isolate one component in the chain of transmission which does not necessarily equate to actual disease (e.g., detection of viral particles vs infectivity vs disease outcomes) [19,20,32]. Results from modelling studies need to be considered alongside epidemiological studies. Previous reviews have highlighted many challenges with studying outbreaks: Li et al [13] mentioned the “most inherent limitation in almost all existing investigations is due to the rapid disappearance of airborne evidence of infection, once the infectious period is over” [13,p.14]. They proposed as a solution “contemporaneous air-sampling and environmental measurements” [13,p.14] in locations during a patient’s illness, which could be extended to locations of high use or occupancy during a pandemic or seasonal epidemics.

### Strengths and Limitations

The strengths of this study include its comprehensiveness and use of methods to avoid bias, such as pre-specification of inclusion/exclusion criteria and involvement of at least two reviewers at all stages. The main limitation stems from the limits of the included reviews. We initially intended to include only systematic reviews that met internationally recognized definitions and methodological expectations. However, we relaxed our criteria given that many reviews did not meet this standard. While most reviews prespecified their research question and conducted a comprehensive search, few conducted study selection and data extraction in duplicate as recommended to avoid bias, and very few assessed the methodological quality or risk of bias of their included studies which is key to determining the validity and certainty of available evidence. We also did not find reviews of all HVAC design features, for example, none of the included reviews examined UVGI (though a recent narrative review has been published in the context of COVID-19 [48]) and only a small number of studies across the reviews examined filtration.

### Implications

The findings of this overview provide several implications for public health measures to mitigate spread of viral transmission in buildings. First, ventilation rates and air flow patterns have been shown to associate with virus transmission. Second, humidity and temperature associate with virus survival. Third, filtration can be effective in removing pathogens if the filter rating is commensurate with the size of the particles of interest [19]. Reviews also mentioned the importance of regular maintenance of HVAC systems and features to ensure optimal functioning. Across the reviews, there was a clearly stated need for more methodologically rigorous inter-disciplinary research with a specific focus on quantifying minimum specifications for HVAC features. While one review did not find sufficient evidence of HVAC and airborne transmission specific to SARS-CoV-2, authors did advise (based on evidence for MERS-CoV and SARS-CoV-1) that attention be given to design and management of HVAC systems as a precautionary measure until further evidence indicates otherwise [21].

## Conclusions

Airborne transmission is now recognized as a route of transmission for different viruses including coronaviruses and specifically SARS-CoV-2 which has been the source of immense global impacts in terms of morbidity, mortality, and peripheral effects of pandemic restrictions. HVAC systems and their specific features have the potential to mitigate transmission in the built environment: there is evidence that ventilation rates, airflow patterns, humidity, temperature, and filtration can influence virus transmission. Enhancing HVAC systems in the built environment (including schools, office buildings, commercial spaces, recreation centers, transport vehicles, etc.) could have important implications for the current pandemic as well as seasonal epidemics and other diseases and impacts that are associated with general indoor air quality. These measures will be of utmost relevance to countries that experience cooler climates and where people spend an inordinate amount of time (80-90%) indoors. Moreover, mitigation strategies that do not rely on human behaviour and result in other (e.g., social) consequences will be more sustainable [21]. This overview synthesized seven previous reviews that included 47 studies examining HVAC design features and their effects on airborne transmission of viruses, serving as a starting point for future systematic reviews and identifying priorities for primary research.

## Data Availability

No additional data available. All data are available in the manuscript and the supplemental material.

## Acknowledgements

We thank Tara Landry and Alison Henry for conducting the peer review of the unfiltered search strategies. We thank Liz Dennett for providing the filter for systematic reviews for Ovid MEDLINE.

## Abbreviations

ACH: air changes per hour
COVID-19: coronavirus disease 2019
HEPA: high efficiency particulate air
HVAC: heating, ventilation and air conditioning
MERS-CoV: Middle East respiratory syndrome coronavirus
MERV: minimum efficiency reporting value
RH: relative humidity
SARS-CoV-1: severe acute respiratory syndrome coronavirus 1
SARS-CoV-2: severe acute respiratory syndrome coronavirus 2
UV: ultraviolet
UVGI: ultraviolet germicidal irradiation
WHO: World Health Organization

---

**Multimedia Appendix 1.**
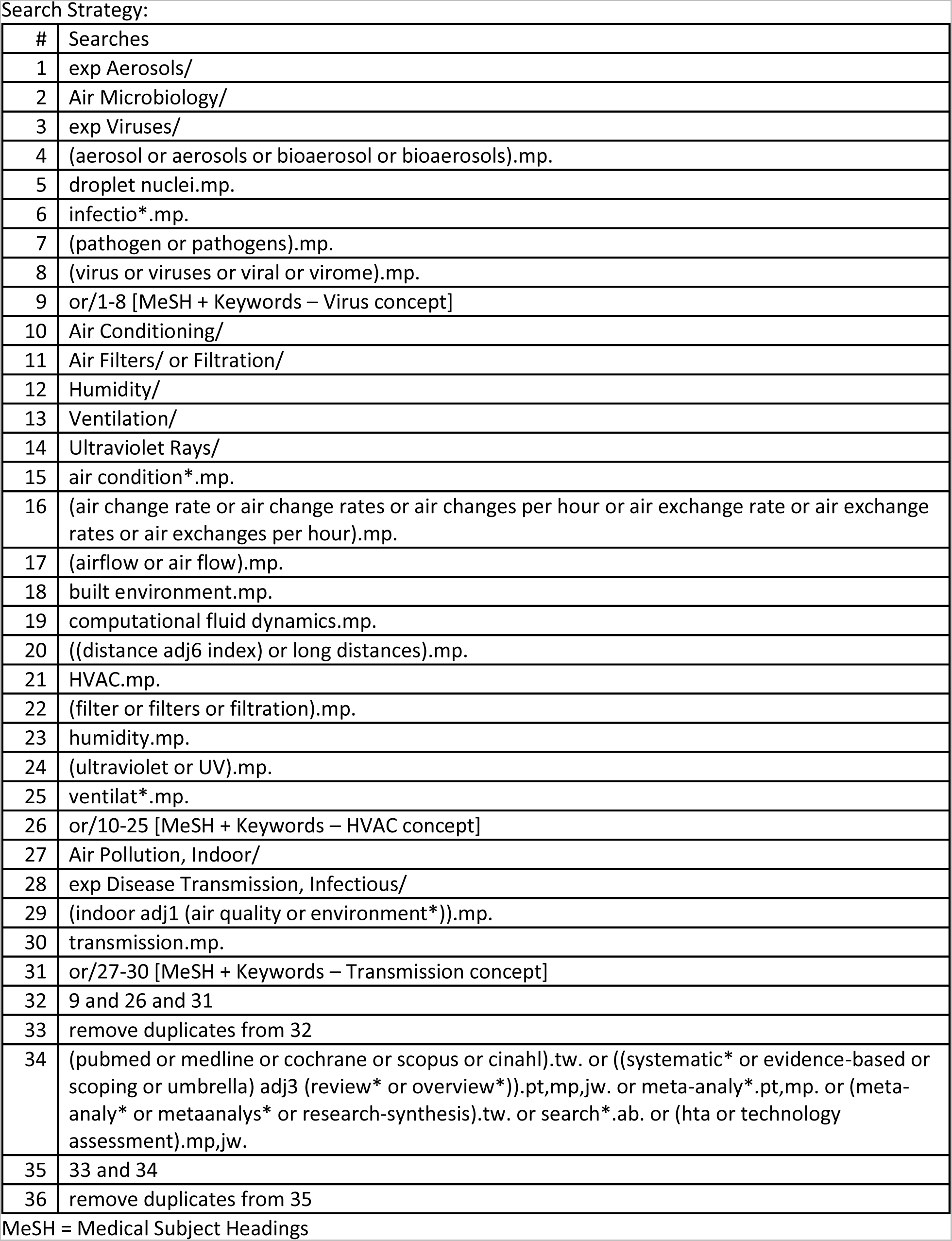
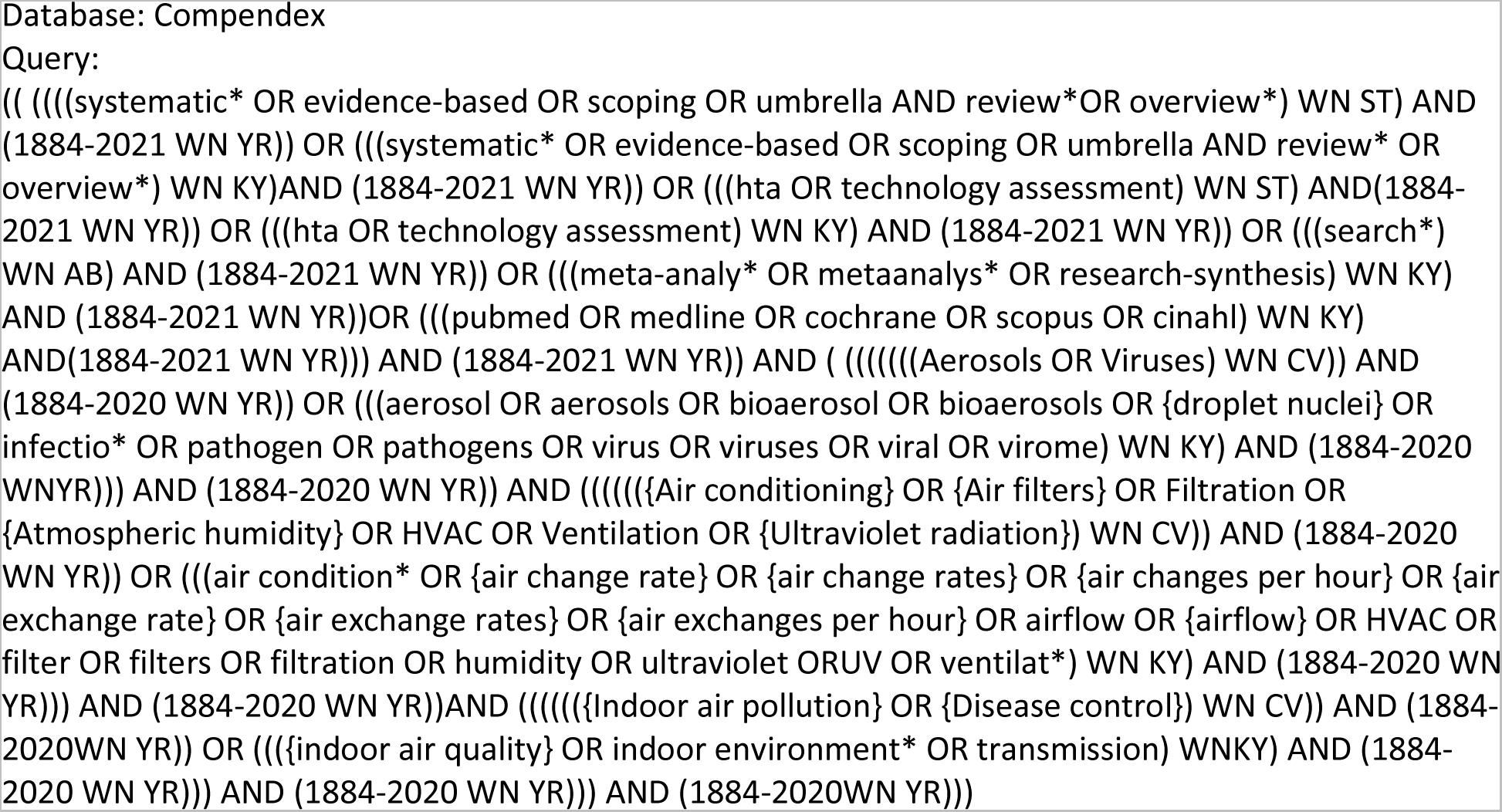
Search Strategies for Ovid MEDLINE and Compendex. Database: Ovid MEDLINE(R) ALL 1946 to Present

**Multimedia Appendix 2.** Inclusion and exclusion criteria for overview of reviews [16].

| Item | Inclusion criteria | Exclusion criteria |
| --- | --- | --- |
| Agent | • Viruses<br>• Aerosols<br>• Bioaerosols<br>• Droplet nuclei<br>• Pathogens<br>*We planned a staged process: if we identify reviews specific to viruses, we will not include reviews not specific to virus.* |  |
| HVAC / Mechanical ventilation | Mechanical ventilation or HVAC overall, or specific to one or more of the following HVAC design features:<br>• Ventilation (ventilation rate, air changes per hour, air exchange, airflow pattern, pressurization)<br>• Filtration (air filtration, filter type, MERV rating, filter age and/or use, pressure drop, holding capacity, replacement, change frequency)<br>• Ultraviolet germicidal irradiation (UVGI; power, dose, uniformity of dose, flow rate, bioaerosol inactivation efficiency, location)<br>• Humidity or relative humidity | • Transmission unrelated to ventilation<br>• Examines only natural ventilation |
| Setting | • Office buildings<br>• Public buildings (e.g., schools, day cares)<br>• Residential buildings<br>• Hospitals and other healthcare facilities (e.g., clinics)<br>• Transport vehicles (e.g., aircraft) or hubs (e.g., airports) | • Outdoor settings<br>• Indoor settings with natural ventilation |
| Outcomes | Quantitative data evaluating the correlation or association between virus transmission and mechanical ventilation or specific HVAC features | Qualitative data |
| Study design | Systematic review, i.e., use of “explicit, systematic methods...aimed at minimizing bias” to “identify, appraise and synthesize all the empirical evidence that meets pre-specified eligibility criteria to answer a specific research question”. (https://www.cochranelibrary.com/about/about-cochrane-reviews)<br>*We planned a staged process: we would include reviews that are not described as systematic if they searched at least 2 databases and/or were specific to viruses and met all other inclusion criteria.* | • Primary studies<br>• Commentaries, opinion pieces |
| Language | English | Non-English |
| Year | No restrictions |  |
| Publication status | Published and/or peer-reviewed | Unpublished and/or not peer-reviewed |

HVAC = heating, ventilation, and air conditioning; MERV = minimum efficiency reporting value; UVGI = ultraviolet germicidal irradiation

**Multimedia Appendix 3.** Publications excluded at full text screening with reasons.

| First author, year | Citation | Reasons for exclusion |
| --- | --- | --- |
| Abd El-Wahab 2020 | Abd El-Wahab EW, Eassa SM, Metwally M, et al. SARS-CoV-2 transmission channels: a review of the literature. *MEDICC Rev* 2020;22:51-69. | Study design: not a systematic review (narrative review of transmission modes and prevention and control strategies) |
| Ahlawat 2020 | Ahlawat A, Wiedensohler A, Mishra SK. An overview on the role of relative humidity in airborne transmission of SARS-CoV-2 in indoor environments. *Aerosol Air Qual Res* 2020;20:1856-61. | Study design: not a systematic review (narrative review of relative humidity and airborne transmission of SARS-CoV-2 in indoor environments) |
| Bing 2018 | Bing-Yuan, Zhang Y-H, Leung NHL, Cowling BJ, Yang Z-F. Role of viral bioaerosols in nosocomial infections and measures for prevention and control. *J Aerosol Sci* 2018;117:200-11. | Study design: not a systematic review (narrative review of viral bioaerosols and nosocomial infections in healthcare settings) |
| Birgand 2020 | Birgand G, Peiffer-Smadja N, Fournier S, et al. Assessment of air contamination by SARS-CoV-2 in hospital settings. *JAMA Netw Open* 2020;3:e2033232. | HVAC: no comparison/quantitative data of HVAC design features |
| Brankston 2007 | Brankston G, Gitterman L, Hirji Z, Lemieux C, Gardam M. Transmission of influenza A in human beings. *Lancet Infect Dis* 2007;7:257-65. | HVAC: no HVAC design features (focus on route of influenza A transmission in humans) |
| Browne 2016 | Browne A, St-Onge Ahmad S, Beck CR, Nguyen-Van-Tam JS. The roles of transportation and transportation hubs in the propagation of influenza and coronaviruses: a systematic review. *J Travel Med* 2016;1-7. | HVAC: no HVAC design features (focus on role of transportation and transportation hubs in transmission of influenza and coronaviruses) |
| Carter 2020 | Carter A. Can orthodontic care be safely delivered during the COVID-19 pandemic? Recommendations from a literature review. *Am J Orthod Dentofacial Orthop* 2020;21:66-7. | Study design: not systematic review (commentary) |
| Cho 2019 | Cho E-M, Hong HJ, Park SH, Yoon DK, Goung SJN, Lee CM. Distribution and influencing factors of airborne bacteria in public facilities used by pollution-sensitive population: a meta-analysis. *Inter J Environ Res Public Health* 2019;16:1483. | Study design: not a systematic review (literature review and field study with pooling of data)<br>Agent: bacteria; not specific to virus |
| Comber 2020 | Comber L, Murchu EO, Drummond L, et al. Airborne transmission of SARS-CoV-2 via aerosols. *Rev Med Virol* 2020;e2184. | HVAC: no HVAC design features |
| Edelson 2012 | Edelson PJ. Patterns of measles transmission among airplane travelers. *Travel Med Infect Dis* 2012;10(5-6):230-5. | HVAC: no HVAC design features |
| Emmerich 2013 | Emmerich SJ, Heinzerling D, Choi J, Persily AK. Multizone modeling of strategies to reduce the spread of airborne infectious agents in healthcare facilities. *Build Environ* 2013;60:105-115. | Study design: primary research (modelling/simulation study), not a systematic review |
| Fox 2020 | Fox GJ, Redwood L, Chang V, Ho J. The effectiveness of individual and environmental infection control measures in reducing the transmission of *Mycobacterium tuberculosis*: a systematic review. *Clin Infect Dis* 2020;72:1-12. | Agent: not specific to virus |
| Guo 2019 | Guo W, Cronk R, Scherer E, Oommen R, Brogan J, Sarr M, Bartram J. A systematic scoping review of environmental health conditions in penal institutions. *Int J Hyg Environ Health* 2019;222:790-803. | Agent: not specific to virus<br>HVAC: no association of virus and HVAC design features |
| Hertzberg 2016 | Hertzberg VS, Weiss H. On the 2-row rule for infectious disease transmission on aircraft. *Ann Glob Health* 2016;82:819-23. | Study design: not a systematic review<br>HVAC: no HVAC design features |
| Irwin 2011 | Irwin CK, Yoon KJ, Wang C, Hoff SJ, Zimmerman JJ, Denagamage T, O'Connor AM. Using the systematic review methodology to evaluate factors that influence the persistence of influenza virus in environmental matrices. *Appl Environ Microbiol* 2011;77:1049-60. | HVAC: examines temperature and humidity but not in context of HVAC and built environment |
| Khalefa 2021 | Khalefa MA, Khadabadi NA, Moores TS, et al. Evidence-based review of safe theatre practice during the COVID-19 pandemic beyond personal protective equipment. *Ann R Coll Surg Engl* 2020;103:88-95. | Study design: not systematic review |
| Leitmeyer 2016 | Leitmeyer K, Adlhoch C. Influenza transmission on aircraft: a systematic literature review. *Epidemiol* 2016;26:743-51. | HVAC: no HVAC design features (focus on transmission of influenza aboard aircraft) |
| Medical Advisory Secretariat 2005 | Medical Advisory Secretariat. Air cleaning technologies: an evidence-based analysis. *Ont Health Technol Assess Ser* 2005;5. | HVAC: no HVAC design features (focus on in-room air cleaners, no studies found) |
| Moffa 2019 | Moffa M, Cronk R, Fejfar D, Dancausse S, Padilla LA, Bartram J. A systematic scoping review of environmental health conditions and hygiene behaviors in homeless shelters. *Inter J Hyg Environ Health* 2019;222:335-46. | Agent: not specific to virus |
| Noorimotlagh 2021 | Noorimotlagh Z, Mirzaee SA, Jaafarzadeh N, et al. A systematic review of emerging human coronavirus (SARS-CoV-2) outbreak: focus on disinfection methods, environmental survival, and control and prevention strategies. *Environ Sci Pollut Res* 2021;28:1-15. | HVAC: no HVAC design features (focus on airborne transmission) |
| Perrone 2021 | Perrone G, Giuffrida M, Bellini V, et al. Operating room setup: how to improve health care professionals safety during pandemic COVID-19—A quality improvement study. *J Laparoendosc Adv Surg Tech* 2021;31:85-9. | HVAC: study examines recommendations and guidelines for operating room set-up; does not search for research about HVAC |
| Raeiszadeh 2020 | Raeiszadeh M, Adeli B. A critical review on ultraviolet disinfection systems against COVID-19 outbreak: applicability, validation, and safety considerations. *ACS Photonics* 2020;7:2941-51. | Study design: not a systematic review (narrative review of ultraviolet disinfection systems against COVID-19 outbreak) |
| Rahimi 2021 | Rahimi NR, Fouladi-Fard R, Aali R, et al. Bidirectional association between COVID-19 and the environment: a systematic review. *Environ Res* 2021;194:110692. | HVAC: no HVAC design features (focus on route of transmission) |
| Ramos 2020 | Ramos CCR, Roque JLA, Sarmiento DB, Suarez LEG, Sunio JTP, Tabungar KIB, Tengco GSC, Rio PC, Hilario AL. *Inter J Health Sci* 2020;14:52-65. | HVAC: no HVAC design features (focus on ultraviolet-C environment sterilization in hospitals) |
| Salonen 2016 | Salonen H, Duchaine C, Letourneau V, Mazaheri M, Laitinen S, Clifford S, Mikkola R, Lappalainen S, Reijula K, Morawska L. Endotoxin levels and contribution factors of endotoxins in resident, school, and office environments – A review. *Atmospheric Environ* 2016;42:360-69. | Study design: not a systematic review<br>Agent: not specific to virus |
| Saran 2020 | Saran S, Gurjar M, Baronia A, Sivapurapu V, Ghosh PS, Raju GM, Maurya I. Heating, ventilation and air conditioning (HVAC) in intensive care unit. *Crit Care* 2020;24:194. | Study design: not a systematic review<br>Agent: looks at standards to maintain good air quality, not to remove/manage virus |
| Scheuch 2020 | Scheuch G. Breathing is enough: for the spread of influenza virus and SARS-CoV-2 by breathing only. *J Aerosol Med Pulm Drug Deliv* 2020;33:230-34. | Study design: letter to the editor, not a systematic review<br>HVAC: focus on whether or not airborne transmission, not specifically about HVAC |
| Seto 2015 | Seto WH. Airborne transmission and precautions: facts and myths. *J Hosp Infect* 2015;89:225-8. | Study design: not a systematic review |
| Sharafi 2020 | Sharafi SM, Ebrahimpour K, Nafez A. Environmental disinfection against COVID-19 in different areas of health care facilities: a review. *Rev Environ Health* 2020;aop. | Study design: not a systematic review<br>HVAC: did not examine association of virus transmission and HVAC |
| Spivakovsky 2020 | Spivakovsky S. Which crucial measures do patients need to follow to prevent transmission of COVID-19 in the dental setting? *Evid Based Dent* 2020;21:79. | Study design: not a systematic review (commentary) |
| Stockwell 2019 | Stockwell RE, Ballard EL, O'Rourke P, Knibbs LD, Morawska L, Bell SC. Indoor hospital air and the impact of ventilation on bioaerosols: a systematic review. *J Hosp Infect* 2019;103:175-84. | Agent: not specific to virus |
| Sundell 2017 | Sundell J. Reflections on the history of indoor air science, focusing on the last 50 years. *Indoor Air* 2017;27:708-24. | Study design: not a systematic review<br>HVAC: did not examine association of virus transmission and HVAC |
| Teunis 2010 | Teunis PF, Brienens N, Kretzschmar ME. High infectivity and pathogenicity of influenza A virus via aerosol and droplet transmission. *Epidemics* 2010;2:215-22. | Study design: not a systematic review<br>HVAC: did not examine association of virus transmission and HVAC |
| Turkistani 2020 | Turkistani KA. Precautions and recommendations for orthodontic settings during the COVID-19 outbreak. *Amer J Orthod Dentofacial Orthop* 2020, In press. | Study design: not a systematic review<br>HVAC: general ventilation but not specific HVAC features |
| Tysiak-Mista 2021 | Tysiak-Mista M, Dubiel A, Brzoza K, Burek M, Palkiewicz K. Air disinfection procedures in the dental office during the COVID-19 pandemic. *Med Pr* 2021;72:39-48. | Study design: not a systematic review<br>HVAC: did not examine association of virus transmission and HVAC |
| Wang 2020 | Wang K, Wu J, Mei W, Wang X. Research status on particulate reduction technology in livestock houses. *Nongye Gongcheng Xuebao/Transactions of the Chinese Society of Agricultural Engineering* 2020;36:204-12. | Language of publication: non-English |
| Wu 2018 | Wu Y, Rong J, Luhung I. Influence of air conditioning and mechanical ventilation (ACMV) systems on indoor microbial aerosols. *Chin Sci Bull* 2018;63:920-30. | Language of publication: non-English |
| Zimmer 2019 | Zimmer C, Leuba SI, Cohen T, Yaesoubi R. Accurate quantification of uncertainty in epidemic parameter estimates and predictions using stochastic compartmental models. *Stat Methods Med Res* 2019;28:3591-608. | Study design: not a systematic review |

**Multimedia Appendix 4.**
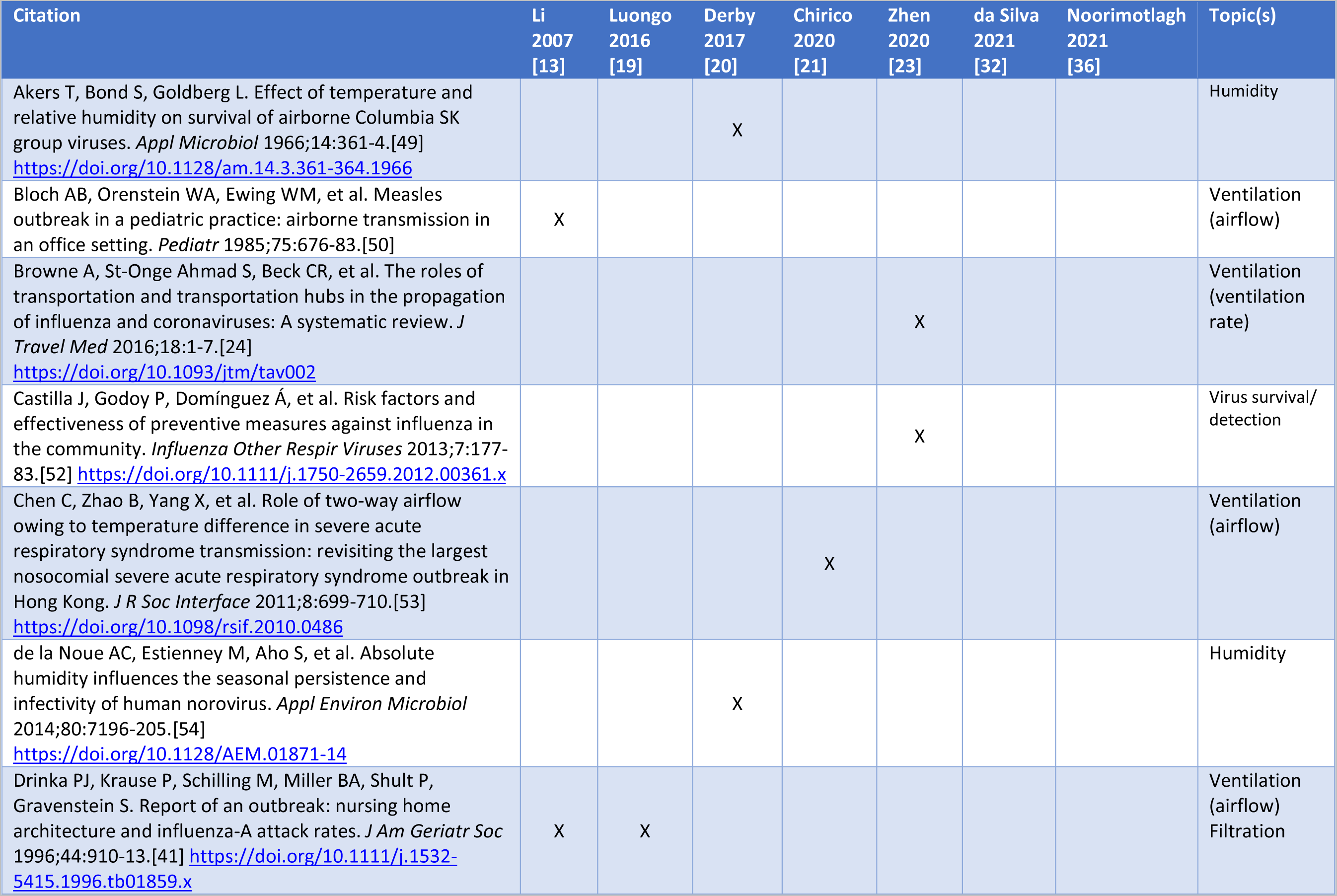
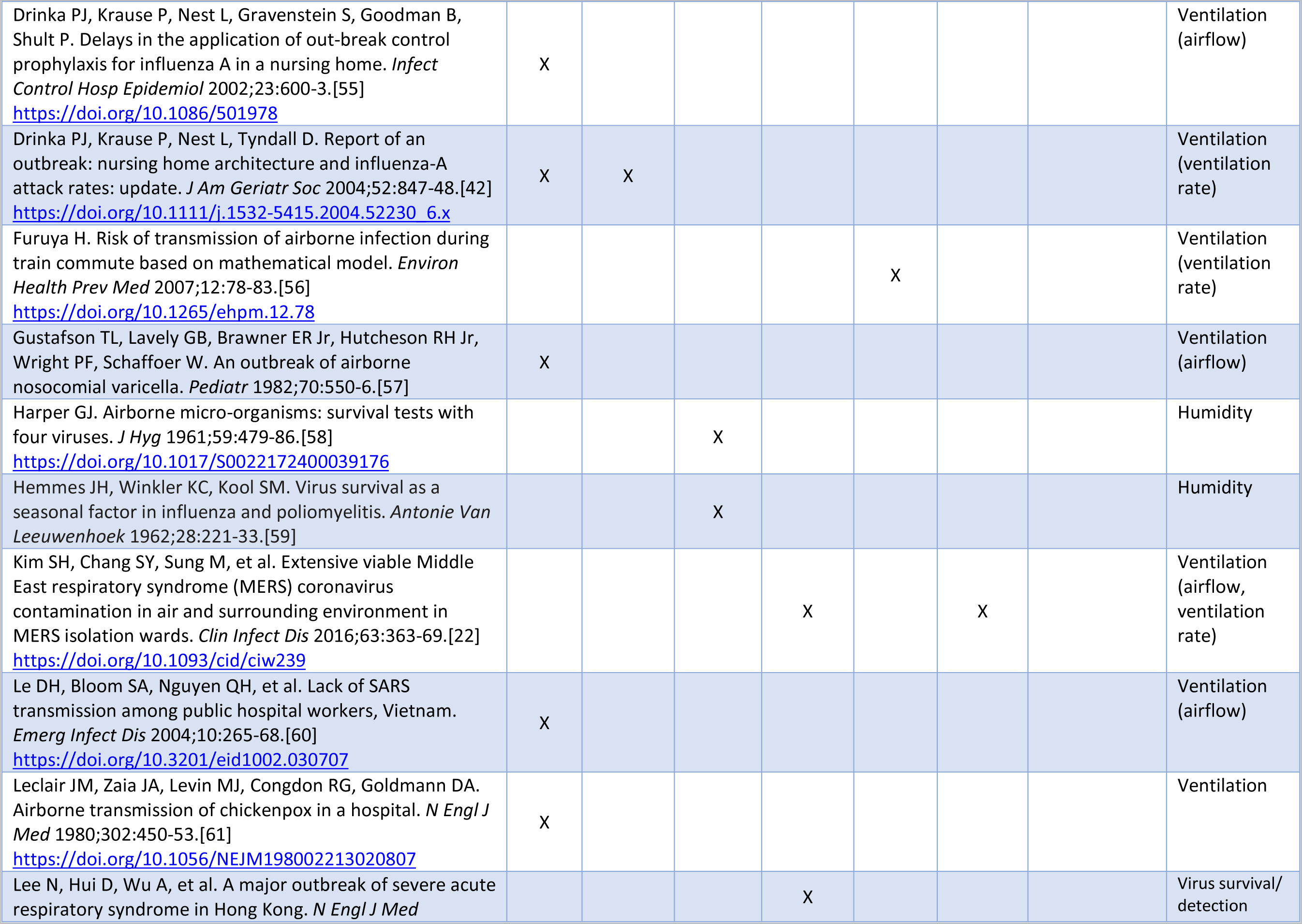
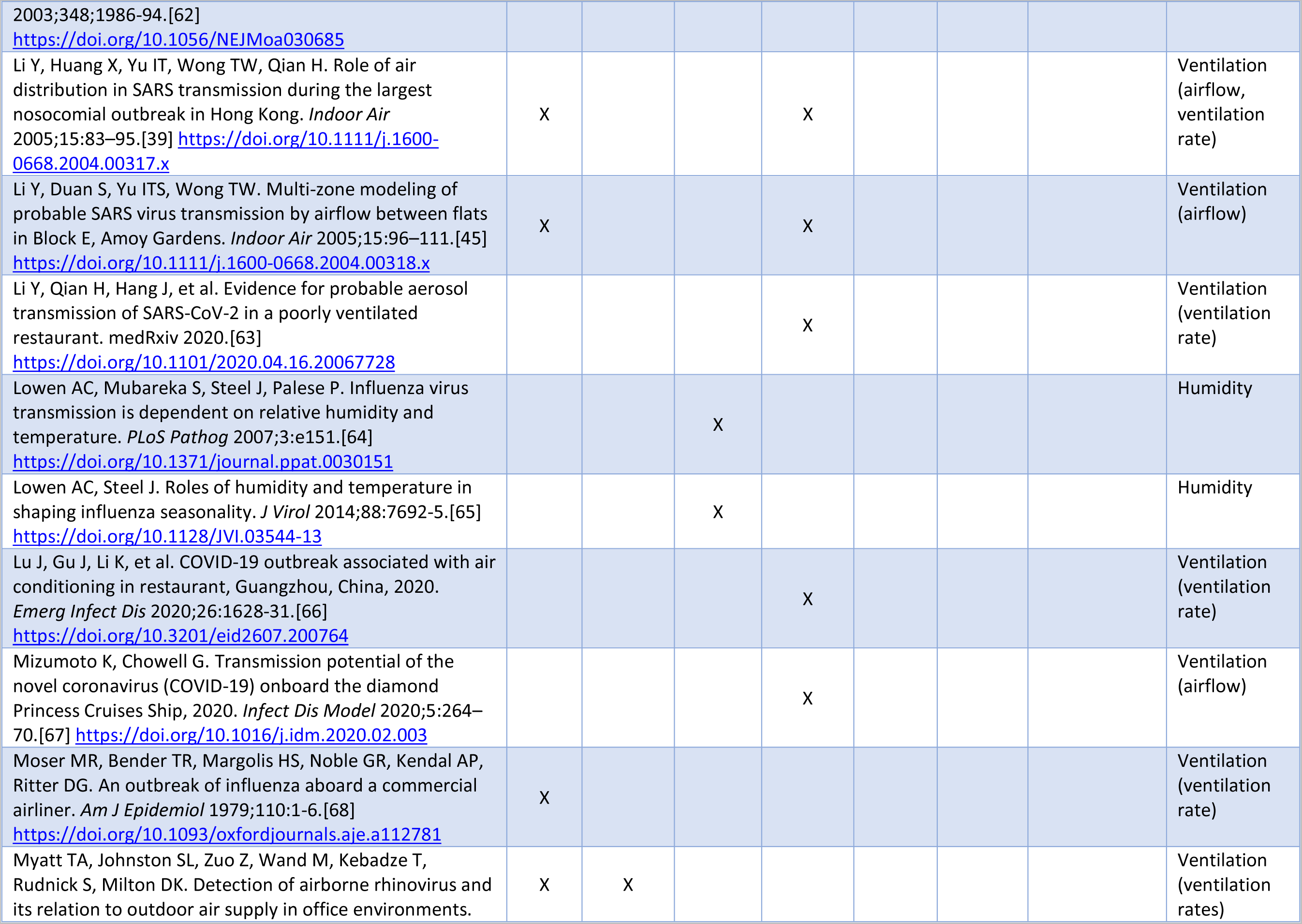
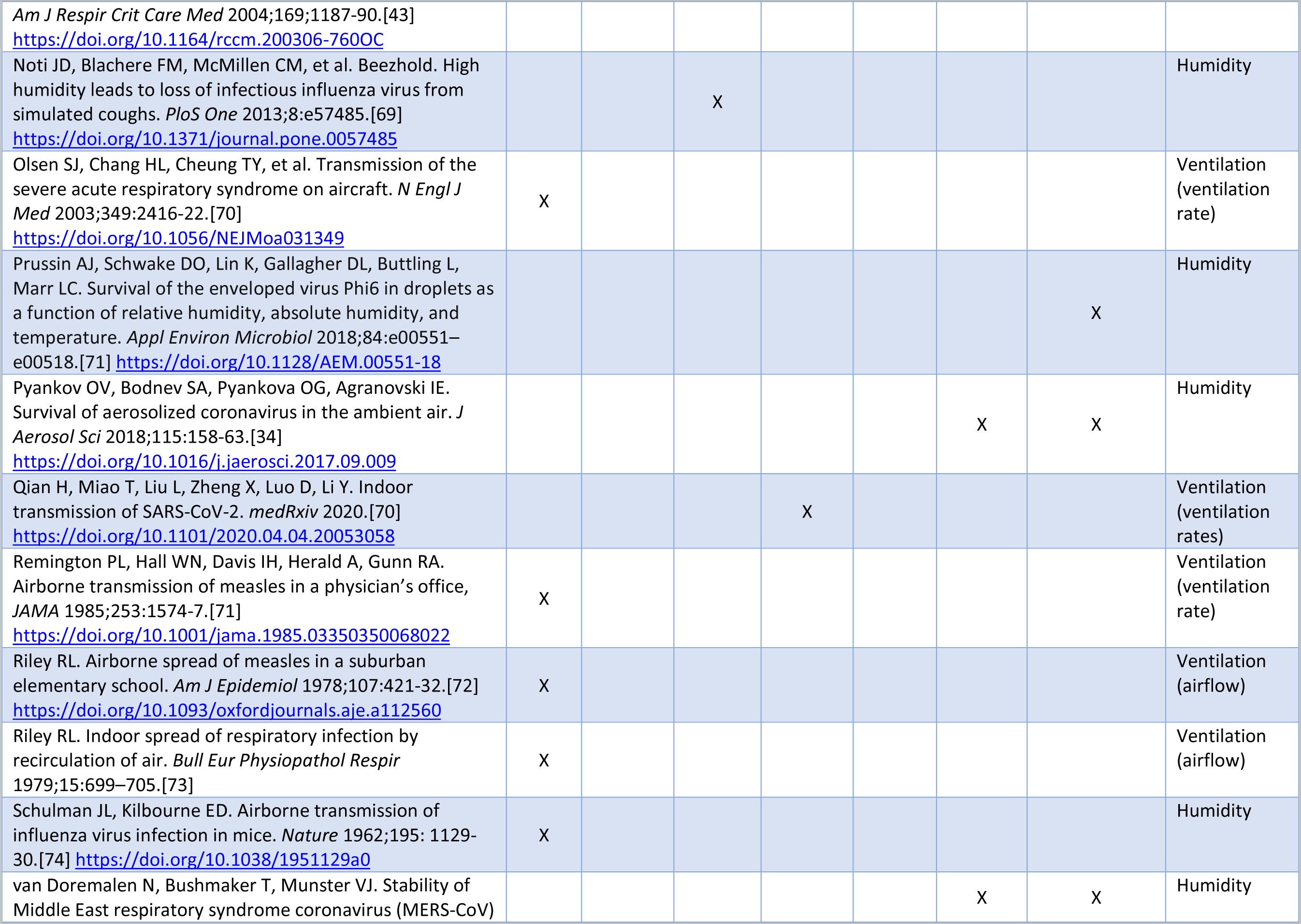
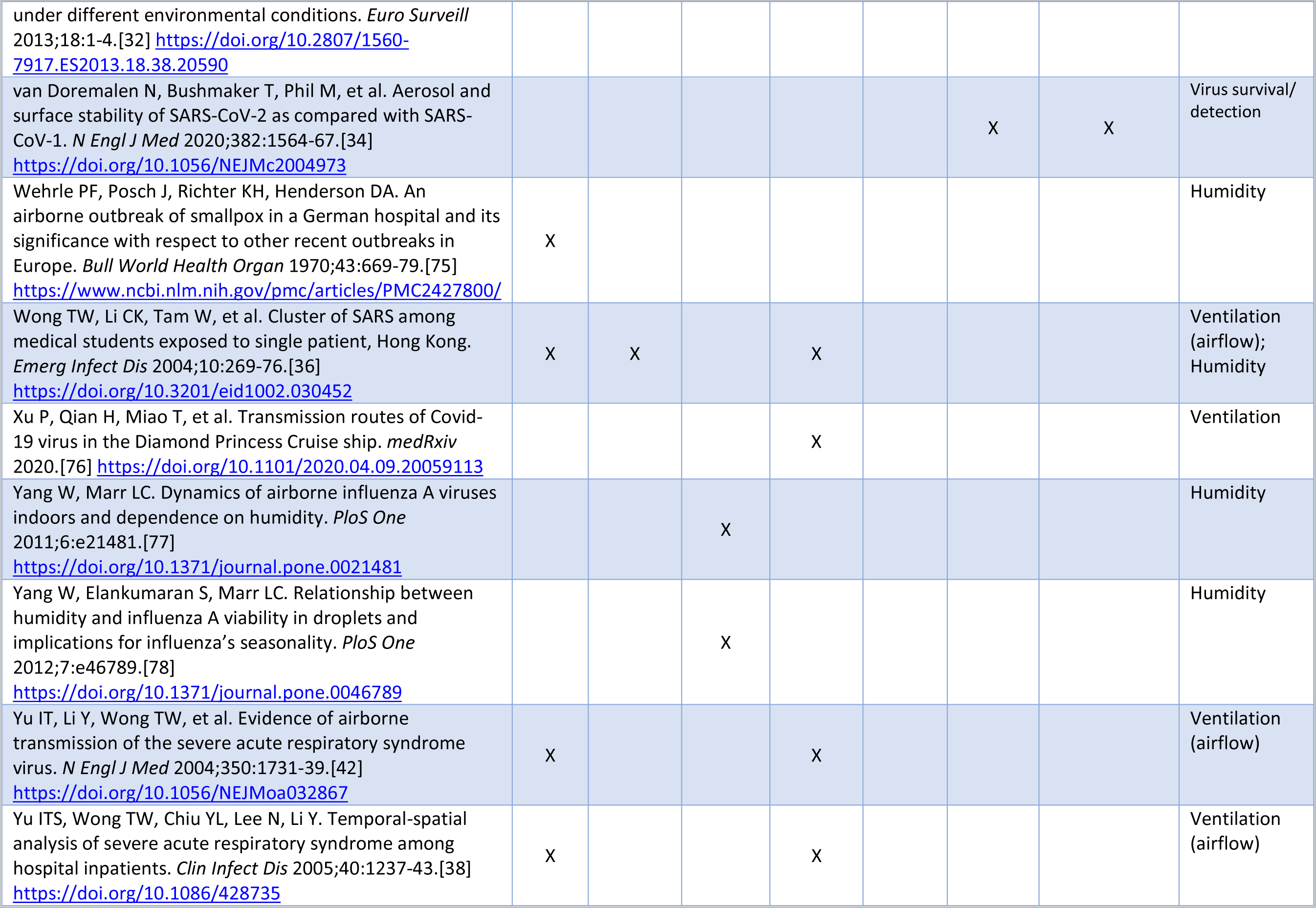
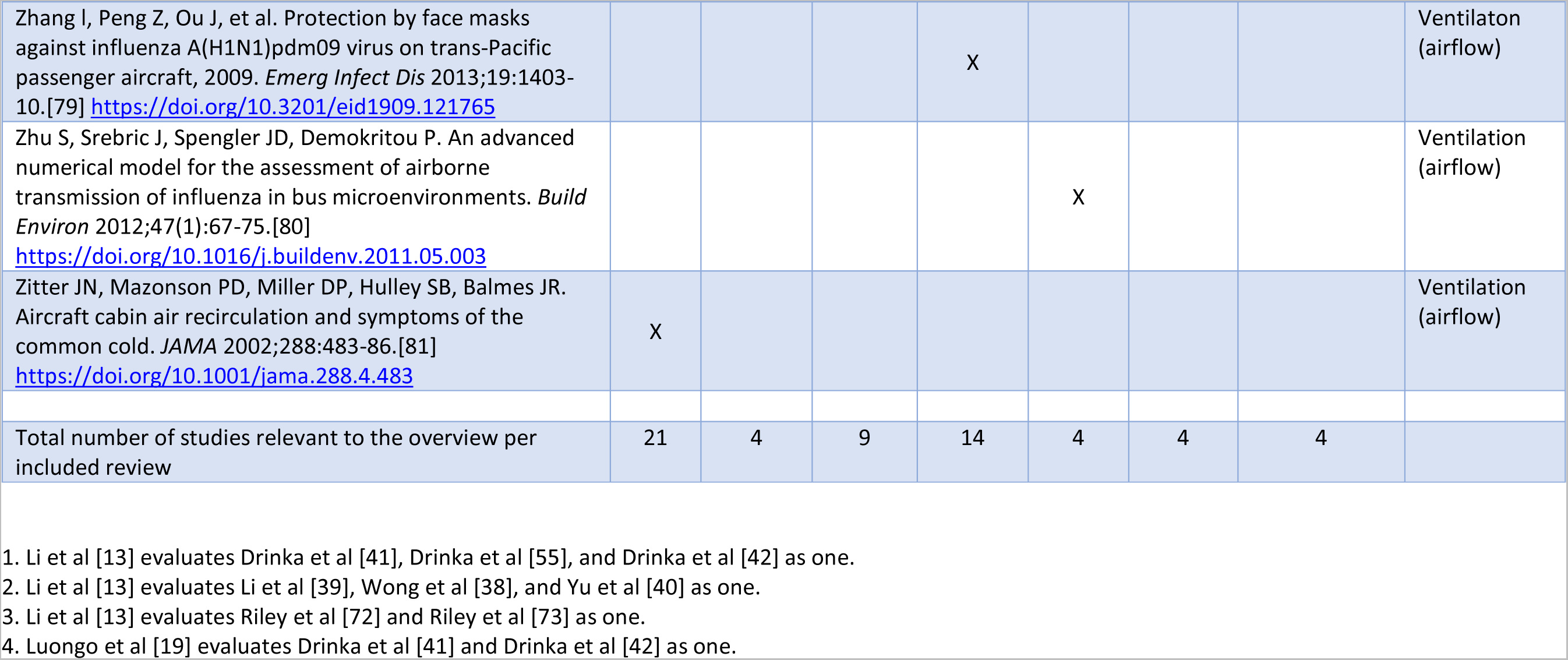
Relevant studies from included reviews that are pertinent to overview research question (With Full Citations)

## References

1. World Health Organization (WHO). WHO Director-General’s opening remarks at the media briefing on COVID-19 - 11 March 2020. [cited 2021 April 4]. Available from: https://www.who.int/director-general/speeches/detail/who-director-general-s-opening-remarks-at-the-media-briefing-on-covid-19 11-march-2020.

2. World Health Organization (WHO). WHO Coronavirus (COVID-19) Dashboard [Internet]. [updated 2022 February 3; cited 2022 February 3]. Available from: https://covid19.who.int

3. COVID-19 transmission–up in the air. Lancet Resp Med 2020;8:1159. https://doi.org/10.1016/S2213-2600(20)30514-2

4. Morawska L, Milton DK. It is time to address airborne transmission of coronavirus disease 2019 (COVID-19). Clin Infect Dis 2020;71:2311 13. https://doi.org/10.1093/cid/ciaa939

5. Baraniuk C. Covid-19: What do we know about airborne transmission of SARS-CoV-2? BMJ 2021;373:n1030. https://doi.org/10.1136/bmj.n1030

6. Tang JW, Marr LC, Li Y, Dancer SJ. Covid19 has redefined airborne transmission. BMJ 2021;373:n913. https://doi.org/10.1136/bmj.n913

7. Comber L, Murchu EO, Drummond L, et al. Airborne transmission of SARS-CoV-2 via aerosols. Rev Med Virol 2020;31:e2184. https://doi.org/10.1002/rmv.2184

8. Rahimi NR, Fouladi-Fard R, Aali R, et al. Bidirectional association between COVID-19 and the environment: a systematic review. Environ Res 2021;194:110692. https://doi.org/0.1016/j.envres.2020.110692.

9. Noorimotlagh Z, Jaafarzadeh N, Martinez SS, Mirzaee SA. A systematic review of possible airborne transmission of the COVID-19 virus (SARS-CoV-2) in the indoor air environment. Environ Res. 2021;193:110612. doi: 10.1016/j.envres.2020.110612

10. Government of Canada. COVID-19: main mode of transmission [Internet]. [updated 2021 March 12; cited 2021 June 18]. https://www.canada.ca/en/public-health/services/diseases/2019-novel-coronavirus-infection/health-professionals/main-modes-transmission.html

11. Brankston G, Gitterman L, Hirji Z, Lemieux C, Gardam M. Transmission of influenza A in human beings. Lancet Infect Dis 2007;7:257–65. https://doi.org/10.1016/S1473-3099(07)70029-4

12. Stockwell RE, Ballard EL, O’Rourke P, Knibbs LD, Morawska L, Bell SC. Indoor hospital air and the impact of ventilation on bioaerosols: a systematic review. J Hosp Infect 2019;103:175e184. https://doi.org/10.1016/j.jhin.2019.06.016

13. Li Y, Leung GM, Tang JW, et al. Role of ventilation in airborne transmission of infectious agents in the built environment – a multidisciplinary systematic review. Indoor Air 2007;17:2–18. https://doi.org/10.1111/j.1600-0668.2006.00445.x

14. Pollock M, Fernandes RM, Becker LA, Pieper D, Hartling L. Chapter V: Overviews of Reviews. In: Higgins JPT, Thomas J, Chandler J, et al., editors. Cochrane handbook for systematic reviews of interventions version 6.1 (updated September 2020). Cochrane, 2020. www.training.cochrane.org/handbook

15. Thornton G, Zhong L, Fleck B, Hartling L. The impact of heating, ventilation and air conditioning (HVAC) design features on the transmission of viruses, including the 2019 novel coronavirus (COVID-19): a systematic review. PROSPERO. 08 July 2020. https://www.crd.york.ac.uk/prospero/display_record.php?ID=CRD42020193968 Accessed July 8, 2020

16. Thornton GM, Fleck BA, Zhong L, Hartling L. The impact of heating, ventilation and air conditioning (HVAC) design features on the transmission of viruses, including the 2019 novel coronavirus (COVID-19): protocol for a systematic review and environmental scan. Open Science Framework. 2020. https://doi.org/10.17605/OSF.IO/Y62V7

17. Shea BJ, Reeves BC, Wells G, et al. AMSTAR 2: a critical appraisal tool for systematic reviews that include randomised or non-randomised studies of healthcare interventions, or both. BMJ 2017;358:j4008. https://doi.org/10.1136/bmj.j4008

18. AMSTAR2 – the new and improved AMSTAR [Internet]. 2017 [cited 2021 June 26]. Available from: https://amstar.ca/Amstar-2.php

19. Luongo JC, Fennelly KP, Keen JA, Zhai ZJ, Jones BW, Miller SL. Role of mechanical ventilation in the airborne transmission of infectious agents in buildings. Indoor Air 2016;26:666–78. https://doi.org/10.1111/ina.12267

20. Derby MM, Hamehkasi M, Eckels S, et al. Update of the scientific evidence for specifying lower limit relative humidity levels for comfort, health, and indoor environmental quality in occupied spaces (RP-1630). Sci Technol Built Environ 2017;23:30–45. https://doi.org/10.1080/23744731.2016.1206430

21. Chirico F, Sacco A, Bragazzi NL, Magnavita N. Can air-conditioning systems contribute to the spread of SARS/MERS/COVID-19 Infection? Insights from a rapid review of the literature. Inter J Environ Res Public Health 2020;17:1–11. https://doi.org/10.3390/ijerph17176052

22. Kim SH, Chang SY, Sung M, et al. Extensive viable Middle East respiratory syndrome (MERS) coronavirus contamination in air and surrounding environment in MERS isolation wards. Clin Infect Dis 2016;63:363–69. https://doi.org/10.1093/cid/ciw239

23. Zhen J, Chan C, Shoonees, Apatu E, Thabane L, Young T. Transmission of respiratory viruses when using public ground transport: a rapid review to inform public health recommendations during the COVID-19 pandemic. S Afr Med J 2020;110:478–83. https://doi.org/10.7196/SAMJ.2020.v110i6.14751

24. Browne A, St-Onge Ahmad S, Beck CR, et al. The roles of transportation and transportation hubs in the propagation of influenza and coronaviruses: A systematic review. J Travel Med 2016;18:1–7. https://doi.org/10.1093/jtm/tav002

25. Government of Canada. Community-based measures to mitigate the spread of coronavirus disease (COVID-19) in Canada [Internet]. [updated 2020 April 3; cited 2020 March 25]. Available from: https://www.canada.ca/en/public-health/services/diseases/2019-novel-coronavirus-infection/health-professionals/public-health-measures-mitigate-covid-19.html

26. European Centre for Disease Prevention and Control (ECDC). Guidelines for the use of non-pharmaceutical measures to delay and mitigate the impact of 2019-nCoV [Internet]. 10 February 2020. Available from: www.ecdc.europa.eu/en/publications-data/guidelines-use-non-pharmaceutical-measures-delay-and-mitigate-impact-2019-ncov (accessed 26 March 2020).

27. GOV.UK. Coronavirus (COVID-19): safer transport guidance for operators [Internet]. [updated 2021 May 17; cited 2021 May 25]. Available from: https://www.gov.uk/government/publications/covid-19-guidance-for-staff-in-the-transport-sector/covid-19-guidance-for-staff-in-the-transport-sector

28. National Academies of Sciences, Engineering, and Medicine. A guide for public transportation pandemic planning and response [Internet]. National Academies Press, 2014 [cited 26 April 2020]. Available from: https://www.nap.edu/read/22414/chapter/1

29. World Health Organization. Infection prevention and control of epidemic- and pandemic-prone acute respiratory infections in health care: WHO guidelines [Internet]. April 2014 [cited 2020 March 25]. Available from: https://www.who.int/csr/bioriskreduction/infection_control/publication/en/

30. World Health Organization. Advice on the use of masks in the community, during home care and in healthcare settings in the context of the novel coronavirus (COVID-19) outbreak [Internet]. 2020 April 6 [cited 2020 April 26]. Available from: www.who.int/publications-detail/advice-on-the-use-of-masks-in-the-community-during-home-care-and-in-healthcare-settings-in-the-context-of-the-novel-coronavirus-(2019-ncov)-outbreak

31. National Department of Health. COVID-19 environmental health guidelines. South Africa, 2020 March 16 [cited 2020 March 25]. Available from: https://j9z5g3w2.stackpathcdn.com/wp-content/uploads/2020/04/COVID-19-ENVIRONMENTAL-HEALTH-GUIDELINE-1-3.pdf

32. Da Silva PG, Nascimento MSJ, Soares RRG, Sousa SIV, Mesquita JR. Airborne spread of infectious SARS-CoV-2: moving forward using lessons from SARS-CoV and MERS-CoV. Sci Total Environ 2021;764:142802. https://doi.org/10.1016/j.scitotenv.2020.142802

33. van Doremalen N, Bushmaker T, Munster VJ. Stability of Middle East respiratory syndrome coronavirus (MERS-CoV) under different environmental conditions. Euro Surveill 2013;18:1–4. https://doi.org/10.2807/1560-7917.ES2013.18.38.20590

34. Pyankov OV, Bodnev SA, Pyankova OG, Agranovski IE. Survival of aerosolized coronavirus in the ambient air. J Aerosol Sci 2018;115:158–63. https://doi.org/10.1016/j.jaerosci.2017.09.009

35. van Doremalen N, Bushmaker T, Phil M, et al. Aerosol and surface stability of SARS-CoV-2 as compared with SARS-CoV-1. N Engl J Med 2020;382:1564–67. https://doi.org/10.1056/NEJMc2004973

36. Noorimatlagh Z, Mirzaee SA, Jaafarzadeh N, Maleki M, Kalvandi G, Karami C. A systematic review of emerging human coronavirus (SARS-CoV-2) outbreak: focus on disinfection methods, environmental survival, and control and prevention strategies. Environ Sci Pollut Res 2021;28:1–15. https://doi.org/10.1007/s11356-020-11060-z

37. Prussin AJ, Schwake DO, Lin K, Gallagher DL, Buttling L, Marr LC. Survival of the enveloped virus Phi6 in droplets as a function of relative humidity, absolute humidity, and temperature. Appl Environ Microbiol 2018;84:e00551–e00518. https://doi.org/10.1128/AEM.00551-18

38. Wong TW, Li CK, Tam W, et al. Cluster of SARS among medical students exposed to single patient, Hong Kong. Emerg Infect Dis 2004;10:269–76. https://doi.org/10.3201/eid1002.030452

39. Li Y, Huang X, Yu IT, Wong TW, Qian H. Role of air distribution in SARS transmission during the largest nosocomial outbreak in Hong Kong. Indoor Air 2005a;15:83–95. https://doi.org/10.1111/j.1600-0668.2004.00317.x

40. Yu ITS, Wong TW, Chiu YL, Lee N, Li Y. Temporal-spatial analysis of severe acute respiratory syndrome among hospital inpatients. Clin Infect Dis 2005;40:1237–43. https://doi.org/10.1086/428735

41. Drinka PJ, Krause P, Schilling M, Miller BA, Shult P, Gravenstein S. Report of an outbreak: nursing home architecture and influenza-A attack rates. J Am Geriatr Soc 1996;44:910–13. https://doi.org/10.1111/j.1532-5415.1996.tb01859.x

42. Drinka PJ, Krause P, Nest L, Tyndall D. Report of an outbreak: nursing home architecture and influenza-A attack rates: update. J Am Geriatr Soc 2004;52:847–48. https://doi.org/10.1111/j.1532-5415.2004.52230_6.x

43. Myatt TA, Johnston SL, Zuo Z, Wand M, Kebadze T, Rudnick S, Milton DK. Detection of airborne rhinovirus and its relation to outdoor air supply in office environments. Am J Respir Crit Care Med 2004;169;1187–90. https://doi.org/10.1164/rccm.200306-760OC

44. Yu IT, Li Y, Wong TW, et al. Evidence of airborne transmission of the severe acute respiratory syndrome virus. N Engl J Med 2004;350:1731–39. https://doi.org/10.1056/NEJMoa032867

45. Li Y, Duan S, Yu ITS, Wong TW. Multi-zone modeling of probable SARS virus transmission by airflow between flats in Block E, Amoy Gardens. Indoor Air 2005b;15:96–111. https://doi.org/10.1111/j.1600-0668.2004.00318.x

46. Menzies D, Fanning A, Yuan L, FitzGerald JM. Hospital ventilation and risk for tuberculous infection in Canadian health care workers. Ann Int Med 2000;133:779–89. https://doi.org/10.7326/0003-4819-133-10-200011210-00010

47. Laminar flow vs turbulent flow. [updated 2021 May 29; cited 26 June 2021]. https://www.archtoolbox.com/materials-systems/hvac/laminarflowvsturbulentflow.html

48. Raeiszadeh M, Adeli B. A critical review on ultraviolet disinfection systems against COVID-19 outbreak: applicability, validation, and safety considerations. ACS Photonics 2020;7:2941–51. https://doi.org/10.1021/acsphotonics.0c01245

49. Akers T, Bond S, Goldberg L. Effect of temperature and relative humidity on survival of airborne Columbia SK group viruses. Appl Microbiol 1966;14:361–4. https://doi.org/10.1128/am.14.3.361-364.1966

50. Bloch AB, Orenstein WA, Ewing WM, et al. Measles outbreak in a pediatric practice: airborne transmission in an office setting. Pediatr 1985;75:676–83.

51. Castilla J, Godoy P, Domínguez Á, et al. Risk factors and effectiveness of preventive measures against influenza in the community. Influenza Other Respir Viruses 2013;7:177–83. https://doi.org/10.1111/j.1750-2659.2012.00361.x

52. Chen C, Zhao B, Yang X, et al. Role of two-way airflow owing to temperature difference in severe acute respiratory syndrome transmission: revisiting the largest nosocomial severe acute respiratory syndrome outbreak in Hong Kong. J R Soc Interface 2011;8:699–710. https://doi.org/10.1098/rsif.2010.0486

53. de la Noue AC, Estienney M, Aho S, et al. Absolute humidity influences the seasonal persistence and infectivity of human norovirus. Appl Environ Microbiol 2014;80:7196– 205. https://doi.org/10.1128/AEM.01871-14

54. Drinka PJ, Krause P, Nest L, Gravenstein S, Goodman B, Shult P. Delays in the application of out-break control prophylaxis for influenza A in a nursing home. Infect Control Hosp Epidemiol 2002;23:600–3. https://doi.org/10.1086/501978

55. Furuya H. Risk of transmission of airborne infection during train commute based on mathematical model. Environ Health Prev Med 2007;12:78–83. https://doi.org/10.1265/ehpm.12.78

56. Gustafson TL, Lavely GB, Brawner ER Jr, Hutcheson RH Jr, Wright PF, Schaffoer W. An outbreak of airborne nosocomial varicella. Pediatr 1982;70:550–6.

57. Harper GJ. Airborne micro-organisms: survival tests with four viruses. J Hyg 1961;59:479–86. https://doi.org/10.1017/S0022172400039176

58. Hemmes JH, Winkler KC, Kool SM. Virus survival as a seasonal factor in influenza and poliomyelitis. Antonie Van Leeuwenhoek 1962;28:221–33.

59. Le DH, Bloom SA, Nguyen QH, et al. Lack of SARS transmission among public hospital workers, Vietnam. Emerg Infect Dis 2004;10:265–68. https://doi.org/10.3201/eid1002.030707

60. Leclair JM, Zaia JA, Levin MJ, Congdon RG, Goldmann DA. Airborne transmission of chickenpox in a hospital. N Engl J Med 1980;302:450–53. https://doi.org/10.1056/NEJM198002213020807

61. Lee N, Hui D, Wu A, et al. A major outbreak of severe acute respiratory syndrome in Hong Kong. N Engl J Med 2003;348;1986–94. https://doi.org/10.1056/NEJMoa030685

62. Li Y, Qian H, Hang J, et al. Evidence for probable aerosol transmission of SARS-CoV-2 in a poorly ventilated restaurant. medRxiv 2020. https://doi.org/10.1101/2020.04.16.20067728

63. Lowen AC, Mubareka S, Steel J, Palese P. Influenza virus transmission is dependent on relative humidity and temperature. PLoS Pathogens 2007;3:e151. https://doi.org/10.1371/journal.ppat.0030151

64. . Lowen AC, Steel J. Roles of humidity and temperature in shaping influenza seasonality. *J* *Virol* 2014;88:7692–5. https://doi.org/10.1128/JVI.03544-13

65. Lu J, Gu J, Li K, et al. COVID-19 outbreak associated with air conditioning in restaurant, Guangzhou, China, 2020. Emerg Infect Dis 2020;26:1628–31. https://doi.org/10.3201/eid2607.200764

66. Mizumoto K, Chowell G. Transmission potential of the novel coronavirus (COVID-19) onboard the diamond Princess Cruises Ship, 2020. Infect Dis Model 2020;5:264–70. https://doi.org/10.1016/j.idm.2020.02.003

67. Moser MR, Bender TR, Margolis HS, Noble GR, Kendal AP, Ritter DG. An outbreak of influenza aboard a commercial airliner. Am J Epidemiol 1979;110:1–6. https://doi.org/10.1093/oxfordjournals.aje.a112781

68. Noti JD, Blachere FM, McMillen CM, et al. Beezhold. High humidity leads to loss of infectious influenza virus from simulated coughs. PloS One 2013;8:e57485. https://doi.org/10.1371/journal.pone.0057485

69. Olsen SJ, Chang HL, Cheung TY, et al. Transmission of the severe acute respiratory syndrome on aircraft. N Engl J Med 2003;349:2416–22. https://doi.org/10.1056/NEJMoa031349

70. Qian H, Miao T, Liu L, Zheng X, Luo D, Li Y. Indoor transmission of SARS-CoV-2. medRxiv 2020. https://doi.org/10.1101/2020.04.04.20053058

71. Remington PL, Hall WN, Davis IH, Herald A, Gunn RA. Airborne transmission of measles in a physician’s office, JAMA 1985;253:1574–7. https://doi.org/10.1001/jama.1985.03350350068022

72. Riley RL. Airborne spread of measles in a suburban elementary school. Am J Epidemiol 1978;107:421–32. https://doi.org/10.1093/oxfordjournals.aje.a112560

73. Riley RL. Indoor spread of respiratory infection by recirculation of air. Bull Eur Physiopathol Respir 1979;15:699–705.

74. Schulman JL, Kilbourne ED. Airborne transmission of influenza virus infection in mice. Nature 1962;195: 1129–30. https://doi.org/10.1038/1951129a0

75. Wehrle PF, Posch J, Richter KH, Henderson DA. An airborne outbreak of smallpox in a German hospital and its significance with respect to other recent outbreaks in Europe. Bull World Health Organ 1970;43:669–79. https://www.ncbi.nlm.nih.gov/pmc/articles/PMC2427800/

76. Xu P, Qian H, Miao T, et al. Transmission routes of Covid-19 virus in the Diamond Princess Cruise ship. medRxiv 2020. https://doi.org/10.1101/2020.04.09.20059113

77. Yang W, Marr LC. Dynamics of airborne influenza A viruses indoors and dependence on humidity. PloS One 2011;6:e21481. https://doi.org/10.1371/journal.pone.0021481

78. Yang W, Elankumaran S, Marr LC. Relationship between humidity and influenza A viability in droplets and implications for influenza’s seasonality. PloS One 2012;7:e46789. https://doi.org/10.1371/journal.pone.0046789

79. Zhang l, Peng Z, Ou J, et al. Protection by face masks against influenza A(H1N1)pdm09 virus on trans-Pacific passenger aircraft, 2009. Emerg Infect Dis 2013;19:1403–10. https://doi.org/10.3201/eid1909.121765

80. Zhu S, Srebric J, Spengler JD, Demokritou P. An advanced numerical model for the assessment of airborne transmission of influenza in bus microenvironments. Build Environ 2012;47(1):67–75. https://doi.org/10.1016/j.buildenv.2011.05.003

81. Zitter JN, Mazonson PD, Miller DP, Hulley SB, Balmes JR. Aircraft cabin air recirculation and symptoms of the common cold. JAMA 2002;288:483–86. https://doi.org/10.1001/jama.288.4.483

